# Early alveolar epithelial cell necrosis is a potential driver of COVID-19-induced acute respiratory distress syndrome

**DOI:** 10.1101/2022.01.23.22269723

**Authors:** Kentaro Tojo, Natsuhiro Yamamoto, Nao Tamada, Takahiro Mihara, Miyo Abe, Mototsugu Nishii, Ichiro Takeuchi, Takahisa Goto

## Abstract

Acute respiratory distress syndrome (ARDS) with COVID-19 is aggravated by hyperinflammatory responses even after the peak of viral load has passed; however, its underlying mechanisms remain unclear. In the present study, analysis of the alveolar tissue injury markers and epithelial cell death markers in patients with COVID-19 revealed that COVID-19-induced ARDS was characterized by alveolar epithelial necrosis at an early disease stage. Serum levels of HMGB-1, one of DAMPs released from necrotic cells, were also significantly elevated in these patients. Further analysis using mouse model mimicking COVID-19-induced ARDS showed that the alveolar epithelial cell necrosis involved two forms of programmed necrosis, namely necroptosis and pyroptosis. Finally, the neutralization of HMGB-1 attenuated alveolar tissue injury in the mouse model. Collectively, necrosis, including necroptosis and pyroptosis, is the predominant form of alveolar epithelial cell death at an early disease stage and subsequent release of DAMPs is a potential driver of COVID-19-induced ARDS.

## Introduction

An infection with a novel strain of coronavirus, severe acute respiratory syndrome coronavirus 2 (SARS-CoV-2), causes coronavirus disease 2019 (COVID-19) pneumonia. In the most severe cases, the disease progresses to acute respiratory distress syndrome (ARDS), which is associated with severe alveolar tissue injury^1, 2^. Interestingly, the disease severity is exacerbated by hyperinflammatory responses even after passing the peak of viral load^3, 4^. However, the mechanisms that underlie disease aggravation in COVID-19-induced ARDS remain unclear. We and others have previously reported that alveolar epithelial injury at a very early disease stage is a hallmark of COVID-19-induced ARDS^5, 6^, suggesting that alveolar epithelial injury may be a trigger of subsequent disease progression. Therefore, elucidating the detailed mechanisms by which alveolar epithelial injury occurs in COVID-19-induced ARDS may reveal a therapeutic target that prevents disease aggravation.

The alveolar epithelial injury in ARDS is characterized by cell death, which is divided into necrosis and apoptosis. Moreover, necrosis comprises not only accidental cell death but also several forms of programmed cell deaths^7, 8^. Although previous studies have demonstrated that both alveolar epithelial necrosis and apoptosis are important for the pathogenesis of ARDS^9^, we have recently demonstrated that necrosis is the predominant form of alveolar epithelial cell death in lipopolysaccharide (LPS)-induced experimental ARDS^10^. In contrast to apoptosis, which does not elicit inflammation, necrosis causes the release of damage-associated molecular patterns (DAMPs) such as high mobility group box (HMGB)-1 from dead cells^11–13^. Therefore, it is possible that the alveolar necrosis during early disease stages and subsequent release of DAMPs may drive disease progression in COVID-19-associated ARDS^14–17^.

Here, we assess whether alveolar epithelial cell necrosis and subsequent release of DAMPs aggravate COVID-19-associated ARDS. To determine the alveolar epithelial cell death patterns in COVID-19 patients with or without ARDS, we analyzed the serum levels of full-length (CK18-M65 antigen) and caspase-cleaved (CK18-M30 antigen) cytokeratin 18, which are epithelial total cell death and epithelial apoptosis markers, respectively, in addition to the other several alveolar epithelial and endothelial injury markers. Moreover, we analyzed the levels of CK18-M65 and CK18-M30 in bronchoalveolar lavage fluids (BALF) from COVID-19-induced ARDS. Finally, we investigated the mechanisms underlying alveolar epithelial cell death using the animal model mimicking COVID-19-induced ARDS^18^, and determined whether blockade of HMGB-1, one of DAMPs released from necrotic cells, can attenuate alveolar tissue injury in the animal model.

Some of the preliminary results of the study have been previously published^5^.

## Results

### Circulating alveolar tissue injury markers in COVID-19 ARDS

Forty-eight (18 non-ARDS and 30 ARDS) of the 84 patients hospitalized with COVID-19 during the study period and 18 healthy volunteers matched as closely as possible for age and sex, were included in the analyses of the circulating markers. Characteristics of patients with COVID-19 are presented in Table 1. Patients with ARDS had higher acute physiology and chronic health evaluation–II (APACHE-II) scores, white blood cell counts, C-reactive protein (CRP) levels, D-dimer levels, and lower ratios of partial pressure of arterial oxygen to fraction of inspired oxygen (P/F ratios) and lymphocyte counts than patients without ARDS on admission. Eight patients with ARDS (26.7%) had died, while among those with ARDS, five developed acute kidney injury, with only a small increase in total bilirubin concentration in several patients. Thus, organ dysfunction in most patients was primarily limited to the lungs.

**Table. 1.** Clinical characteristics of ARDS and non-ARDS patients with COVID-19.

|  | Non-ARDS (n = 18) | ARDS (n = 30) | p-Value |
| --- | --- | --- | --- |
| Age (years) | 65 (49–74) | 69 (63–76) | 0.2288 |
| Males/Females | 13/5 | 23/7 | 0.7519 |
| APACHE2 score | 8.0 (6.5–10.3) | 12.5 (9.0–15.0) | *0.0005 |
| P/F ratio at admission | 405.0 (323.1–448.8) | 170.3 (112.8–238.8) | * < 0.0001 |
| Mechanical ventilation use | 0 (0.0%) | 30 (100.0%) | * < 0.0001 |
| In hospital mortality | 0 (0.0%) | 8 (26.7 %) | *0.0182 |
| Laboratory data on admission |  |  |  |
| WBC count (/μL) | 5550 (2,925–7,700) | 7850 (6,175–10,675) | *0.0178 |
| Lymphocyte count (/μL) | 908 (611–1,268) | 469 (273–803) | *0.0040 |
| Platelet count ($\times 10^3/\mu\text{L}$) | 185.5 (123.8–270.0) | 188.0 (138.8–263.5) | 0.7922 |
| D-dimer (μg/mL) | 0.72 (0.00–2.41) | 1.22 (0.99–1.35) | *0.0185 |
| CRP (mg/dL) | 1.04 (3.48–4.21) | 12.32 (7.22–17.22) | * < 0.0001 |
| Creatinine (mg/dL) | 0.84 (0.72–6.76) | 0.81 (0.63–1.49) | 0.3074 |
| Total bilirubin (mg/dL) | 0.55 (0.40–0.73) | 0.50 (0.40–0.90) | 0.8028 |
| Treatment |  |  |  |
| Systemic Corticosteroids | 23 (76.7%) | 4 (22.2%) | *0.0003 |
| Inhaled Corticosteroids | 25 (83.3%) | 11 (61.1%) | 0.1008 |
| Favipiravir | 10 (33.3%) | 8 (44.4%) | 0.5425 |
| Remdesivir | 16 (53.3%) | 1 (5.6%) | *0.0007 |
| Tocilizumab | 4 (13.3%) | 0 (0.0%) | 0.2824 |
| Lopinavir-Ritonavir | 7 (23.3%) | 2 (11.1%) | 0.4511 |
| Past medical history |  |  |  |
| Hypertension | 14 (46.7%) | 7 (11.1%) | 0.7652 |
| Diabetes mellitus | 5 (16.7%) | 7 (11.1%) | > 0.9999 |
| Respiratory diseases | 5 (16.7%) | 3 (16.7%) | > 0.9999 |
| Cardiovascular diseases | 3 (10.0%) | 4 (22.2%) | 0.4002 |
| Liver diseases | 3 (10.0%) | 1 (5.6%) | > 0.9999 |
| Kidney diseases | 4 (13.3%) | 5 (27.8%) | 0.2654 |

Data are presented as count (%) or median (IQR); \*p < 0.05.

We evaluated the circulating levels of three alveolar tissue injury markers: an alveolar epithelial injury marker (sRAGE)^19,20^and an endothelial injury marker (ANG-2)^21, 22^, along with an alveolar permeability indicator (SP-D)^23, 24^. All alveolar tissue injury markers levels after the admission were significantly higher in patients with ARDS versus healthy controls (Fig. 1A–C). However, only sRAGE and SP-D levels of patients with and without ARDS significantly differed (Fig. 1A–C). In patients with ARDS, sRAGE levels were significantly elevated at admission, and gradually decreased thereafter (Fig. 1D, G). Meanwhile, ANG-2 and SP-D levels peaked later (Fig. 1E–G). Collectively, these results agree with prior work that demonstrated that severe alveolar epithelial cell injury at a very early disease stage is a hallmark of COVID-19-induced ARDS^5, 6^.

**Figure 1.**
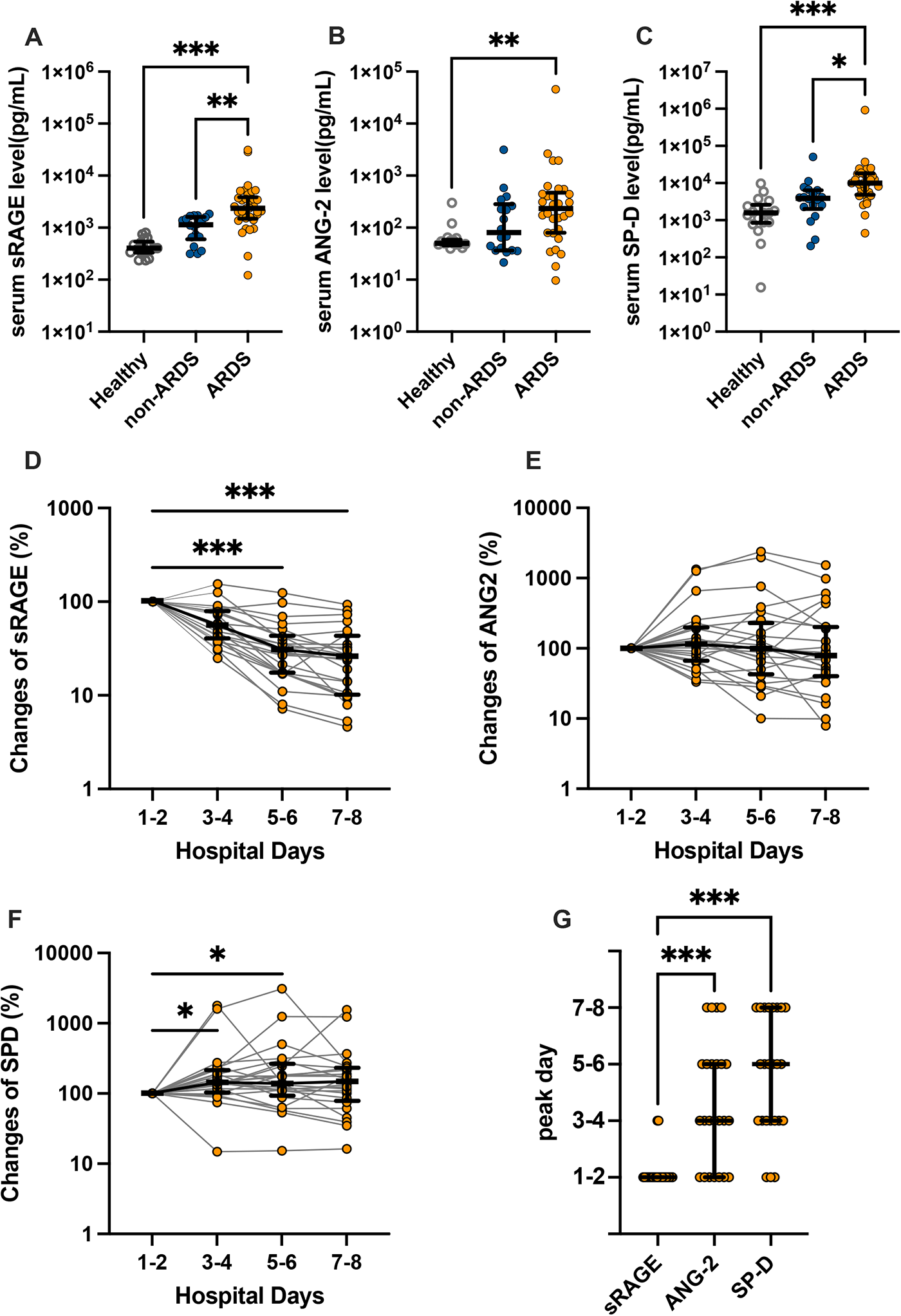
Analysis of serum levels of alveolar tissue injury markers using enzyme linked immunosorbent assays (ELISAs). (A) soluble receptors for advanced glycation end products (sRAGE), (B) angiopoietin (ANG)-2, and (C) surfactant protein (SP)-D levels in the serum of patients with COVID-19 with or without acute respiratory distress syndrome (ARDS) at admission (on the first or second hospital day), and healthy controls are shown. Bidaily temporal changes in (D) sRAGE, (E) ANG-2, and (F) SP-D in sera of COVID-19 patients with ARDS during first 8 days after hospital admission are shown. In cases in which multiple values every 2 days were available, mean values were used. When only a single value was available, the value was used. (G) Days in which concentrations of each alveolar tissue injury marker peaked in COVID-19 patients with ARDS are shown. Values are presented as medians with interquartile ranges. *p < 0.05, **p < 0.01, ***p < 0.0001.

### Epithelial necrosis markers and HMGB-1 are increased in COVID-19 ARDS

Next, the levels of epithelial cell death markers were evaluated to elucidate the predominant form of alveolar epithelial cell death at early disease stage of COVID-19-induced ARDS. The serum levels of CK18-M65 and -M30 antigens were measured to distinguish alveolar necrosis from apoptosis. CK18 is exclusively expressed in epithelial cells and is released upon cell death. The M65 antigen is an indicator of both epithelial cell necrosis and apoptosis. In contrast, the M30 antigen produced after caspase cleavage of CK18 is an indicator apoptotic epithelial cell death^25^. Although CK18 is expressed in all kinds of epithelial cells, the most part of CK18 was presumably derived from alveolar epithelial cells in this cohort because the organ damage was limited almost exclusively to the lungs. Serum levels of both M65 and M30 at admission was positively correlated with disease severity in patients with COVID-19 (Fig. 2A, B), suggesting that both apoptosis and necrosis contribute to alveolar epithelial cell death in COVID-19. The M30/M65 ratio, which is an indicator of fraction of apoptosis among total epithelial cell death, was significantly lower in ARDS patients [median: 31.5 %, IQR: 19.4−43.3] than non-ARDS patients [median: 46.7 %, IQR: 36.6−80.5] or healthy controls [median: 98.9 %, IQR: 83.1−100.0] (Fig. 2C). Moreover, we analyzed the CK18-M30/M65 ratio in BALF, which directly reflects the pulmonary pathology, from six patients with COVID-19 ARDS (Fig. S1). The characteristics of the included patients in the BALF analysis are shown in Table S1. The M30/M65 ratio in the BALF was 27.8% [IQR: 13.3−38.5] (Fig. S1), similar to that of serum samples. Collectively, these results indicate that alveolar epithelial cell death in COVID-19 ARDS is predominantly caused by necrosis. The serum levels of CK18-M65 (Fig. 2D), but not the CK18-M30 levels and the M30/M65 ratios (Fig. 2E, F), at 7 or 8 days after admission were significantly lower than those just after hospital admission.

**Figure 2.**
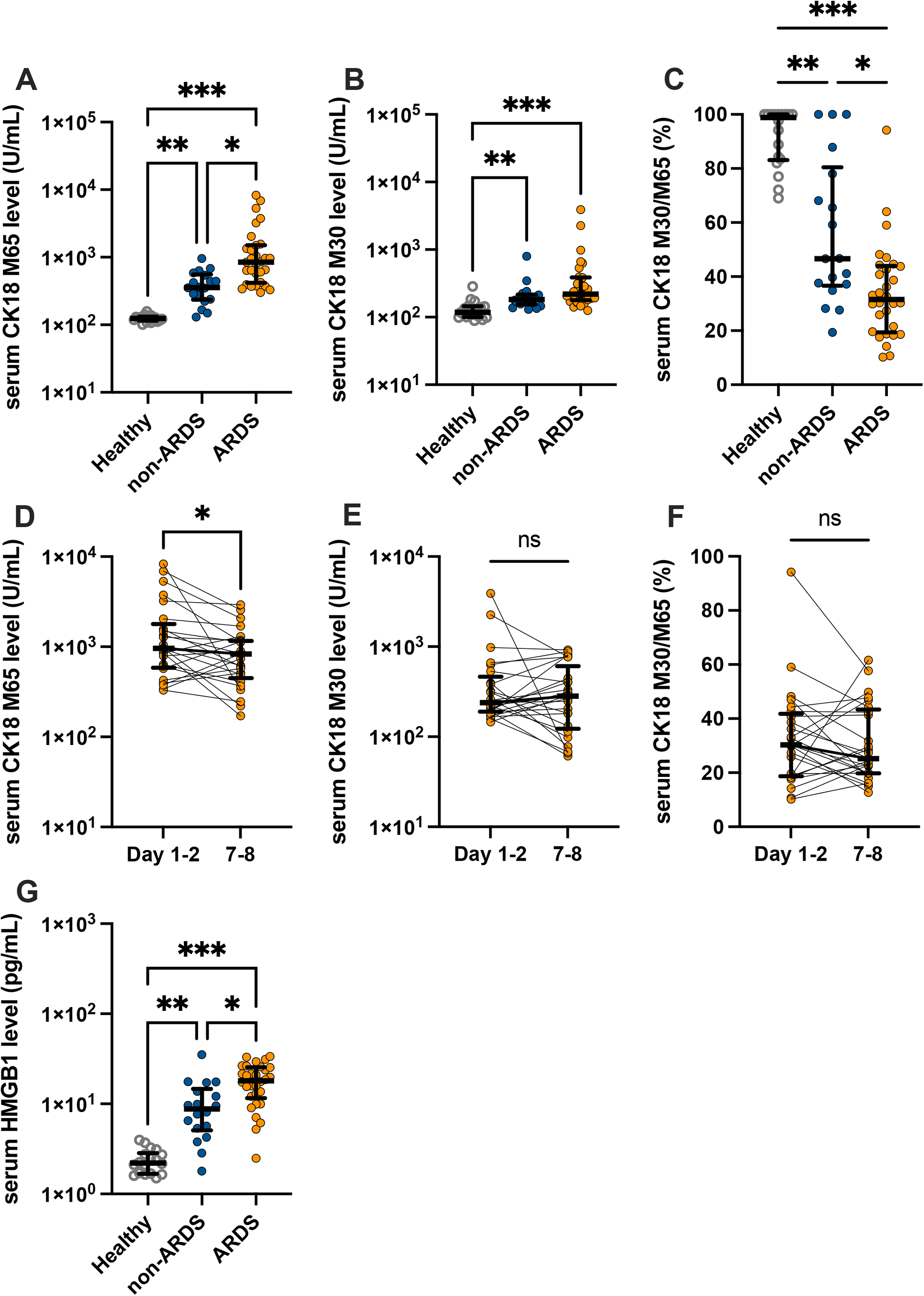
Serum levels of markers of epithelial cell death and high mobility group box (HMGB)-1 in serum samples of COVID-19 patients with or without acute respiratory distress syndrome (ARDS) and healthy controls. Levels of (A) CK18-M65, an epithelial total cell death marker, and (B) CK18-M30, an epithelial apoptosis marker; (C) CK18-M30/M65 ratio, an indicator of the fraction of epithelial cells undergoing apoptosis versus all types of cell death at admission (on the first or second hospital day). Temporal changes in (D) CK18-M65, (E) CK18-M30, and (F) CK18-M30/M65 ratio in sera of COVID-19 patients with ARDS. (G) HMGB-1 levels in serum samples of COVID-19 patients with or without ARDS and healthy controls are shown. Values are presented as medians and interquartile ranges. *p < 0.05, **p < 0.01, ***p < 0.0001.

Necrosis, unlike apoptosis, induces the release of DAMPs and augmentation of inflammation. HMGB-1, a most extensively studied DAMPs mediating organ injury during ARDS or sepsis, is increased in severe COVID-19. We also confirmed that serum levels of HMGB-1 were significantly elevated in ARDS versus non-ARDS patients and healthy controls (Fig. 2G). Moreover, the analysis of correlations among these biomarkers in the serum of COVID-19 patients demonstrated that HMGB-1 levels were most strongly correlated with levels of total epithelial cell death marker, M65 (correlation coefficient = 0.612, p < 0.0001, Fig. S2).

### Intratracheal instillation of SARS-CoV-2 spike proteins combined with poly (I:C) to mice induces lung injury mimicking COVID-19-induced ARDS

The innate immune responses to components of SARS-CoV-2 are principal drivers of inflammation and alveolar tissue injury in COVID-19^26–28^. To elucidate mechanisms underlying alveolar epithelial cell death in COVID-19-induced ARDS, we established animal models of severe and mild COVID-19 by intratracheal instillation with the SARS-CoV-2 spike protein and poly (I:C), a synthetic analog of double-stranded RNA based on previous reports ^18^ and our preliminary experiments (Fig. S3). In the COVID-19 animal model, leukocytes infiltration (Fig. 3A), increased levels of protein, sRAGE, and ANG-2 in BALF (Fig. 3A–D), and lung tissue injury (Fig. 3E) were observed. Moreover, levels of several chemokines and cytokines previously reported to be elevated in COVID-19 patients^29, 30^ were significantly increased in the BALF of animal models of severe COVID-19 versus controls (Fig. 3F).

**Figure 3.**
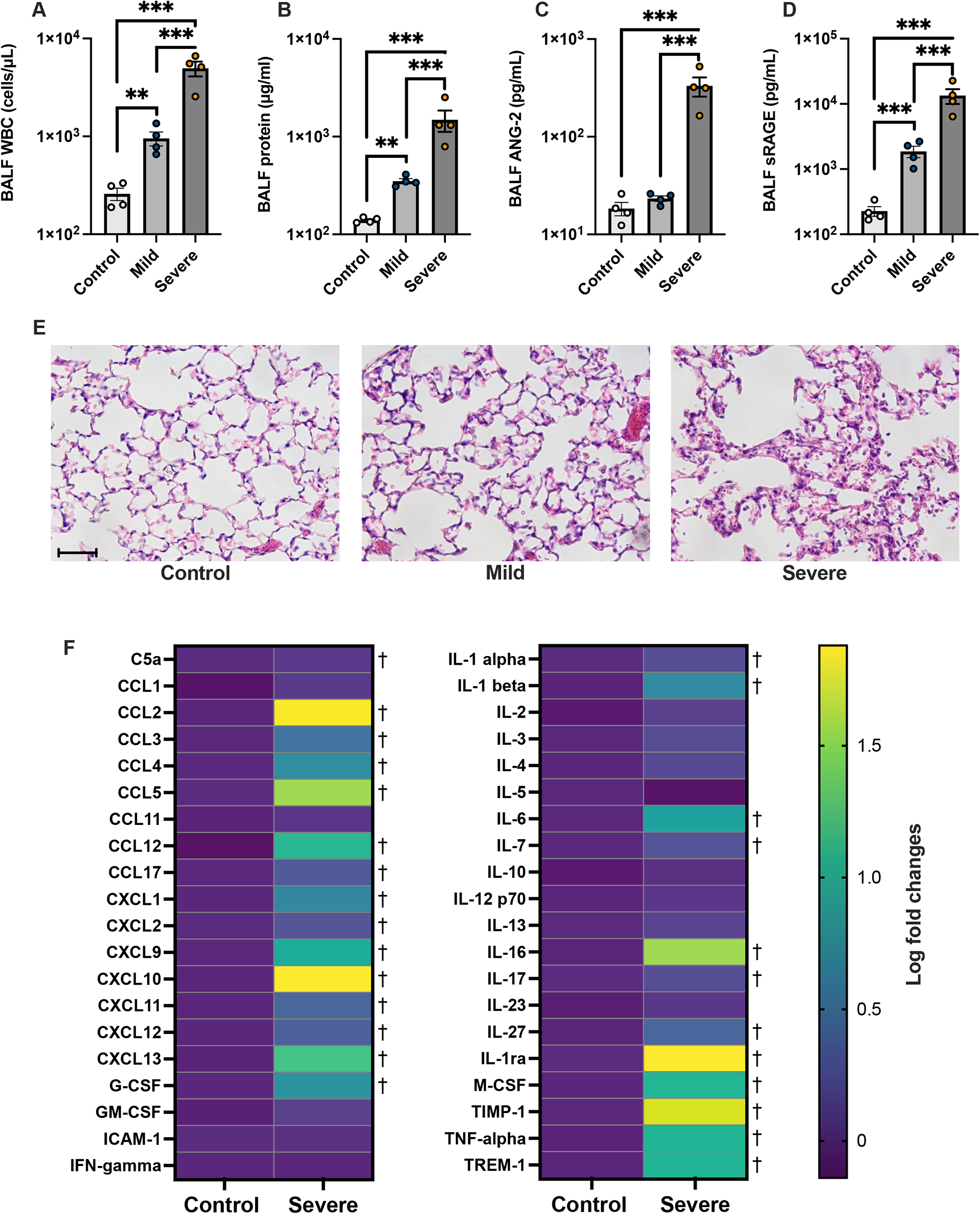
Use of a mouse model of mild and severe COVID-19. (A) White blood cell count, (B) total protein, (C) soluble receptors for advanced glycation end products (sRAGE), and (D) angiopoietin (ANG)-2 levels in bronchoalveolar lavage fluid (BALF) of mouse models of COVID-19 and controls are shown. (E) Representative lung tissue images of sections stained with hematoxylin and eosin are shown. Scale Bar = 50 μm.Values are presented as means ± standard error. *p < 0.05, **p < 0.01, ***p < 0.0001. (F) A heatmap constructed from the comprehensive analysis of cytokine levels in the BALF of mouse models of severe COVID-19 versus control mice. †q-value < 0.05.

We also performed bioinformatic analysis of lung tissue transcriptomes to determine whether the COVID-19 model using SARS-CoV-2 spike protein and poly (I:C) recapitulated the biological responses observed in previously reported mouse models infected with SARS-CoV-2. We identified 3,491 upregulated and 3,174 downregulated differentially expressed genes (DEGs) in our severe COVID-19 model compared with the control (Fig. S4). Furthermore, the gene set enrichment analysis (GSEA)^31^ for REACTOME pathways revealed that immunological and inflammatory pathways were upregulated, while metabolic pathways were downregulated in the severe COVID-19 model (Fig. 4A). We found 6 publicly available lung tissue RNA-seq datasets of mice infected with SARS-CoV-2 from the NCBI Gene Expression Omnibus database (Table S3)^32–37^ and performed comparative analysis for all the datasets including the data obtained from this study. Comparison of differentially expressed genes (DEGs) showed only small overlaps, even when comparisons were performed among the infection models (Table S4). However, reciprocal GSEA^31, 38^ revealed high concordance in upregulated gene sets and modest concordance in downregulated gene sets (Fig.4B). Additionally, we compared the normalized enrichment scores (NESs) of REACTOME pathways among the datasets. The pathways significantly changed in least one dataset were included in this analysis. The patterns of NESs showed high concordance among the groups (Fig. 4C), with a strong correlation between NES in our data and mean NES of the previously reported infection models (correlation coefficient = 0.71, p < 0.001, Fig. 4D). Finally, analysis of the gene expression patterns in several key inflammatory and cell death pathways showed apparent concordances in the gene expressions between the COVID-19 model in the present study and the infection models (Fig. S5). Collectively, bioinformatic analysis demonstrated that our COVID-19 mimicking model recapitulated the biological responses in the mice infected with SARS-CoV-2, at least in the key immunological and cell death pathways.

**Figure 4.**
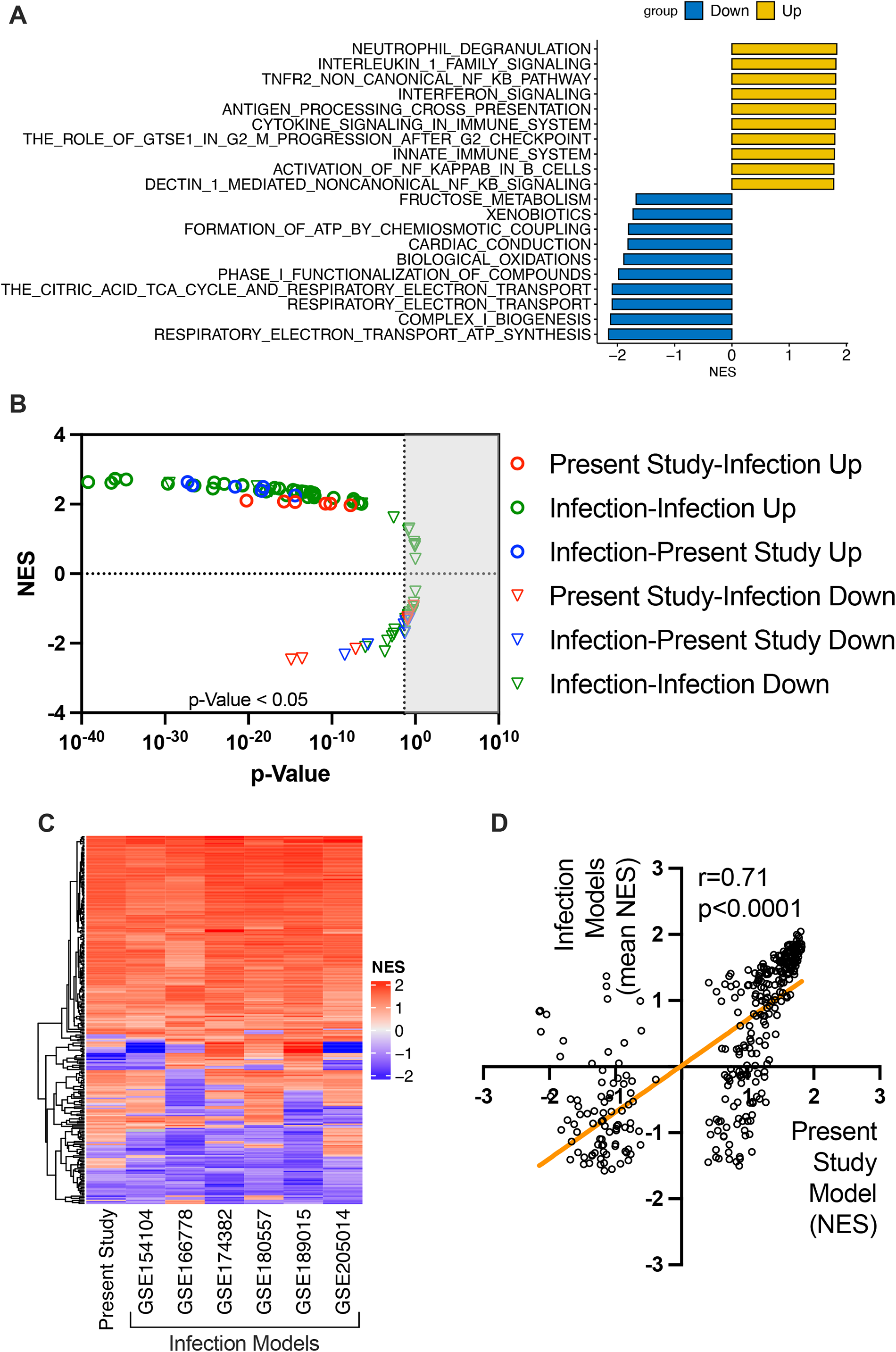
Analysis of lung tissue transcriptomes in the COVID-19 mouse model in the present study and previously reported infection models. (A) Top 10 upregulated and downregulated REACTOME pathways in the severe COVID-19 mouse model in the present study. (B) Reciprocal GSEA (C) Heatmap of NES in each dataset, and (D) correlation between the NES of the severe COVID-19 model in the present study and the mean NES of previously reported infection models. In these analysis (C, D), REACTOME pathways significantly changed at least in one dataset were included.

### Necrosis, including necroptosis and pyroptosis, is a predominant form of alveolar epithelial cell death in the mouse model of severe COVID-19

Levels of both CK18-M30 and total CK18, which is equivalent to CK18-M65, were increased in the BALF of COVID-19 models versus controls (Fig. 5A, B), indicating that both necrosis and apoptosis are involved in alveolar epithelial cell death. CK18-M30/total CK18 ratio, an indicator of apoptosis fraction relative to total epithelial cell death, decreased as lung injury increased in severity, as observed in COVID-19 patients (Fig. 5C). Additionally, the HMGB-1 levels were significantly elevated in severe COVID-19 animal model versus the other two groups (Fig.5D). Taken together, these results demonstrated that the animal model of severe COVID-19 exhibited the same pattern of alveolar epithelial cell death as COVID-19 patients with ARDS.

**Figure 5.**
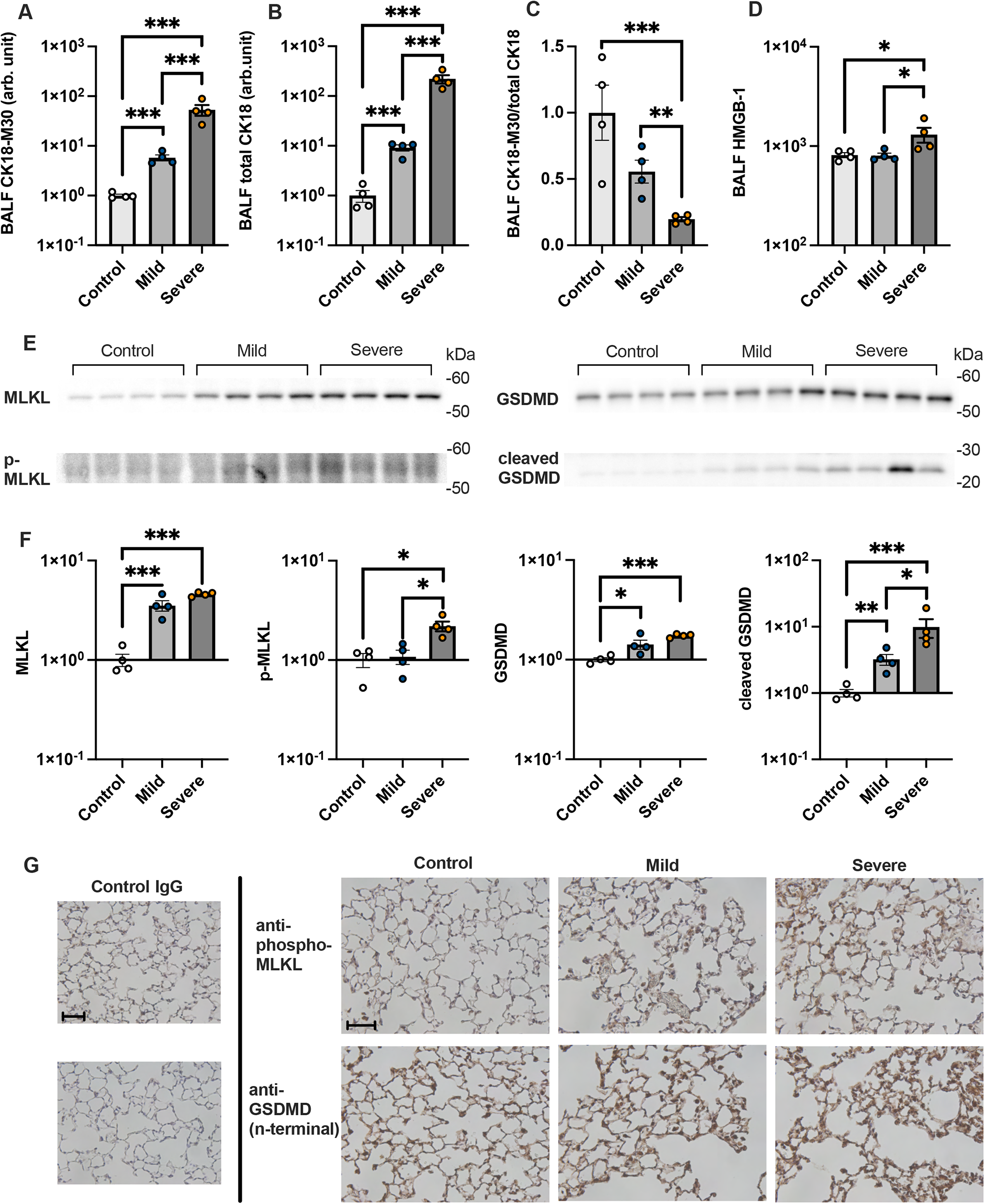
Mechanisms of alveolar epithelial cell death in a COVID-19 mouse model. Levels of (A) CK18-M30 and (B) total CK18in bronchoalveolar lavage fluid (BALF) from a COVID-19 mouse model are shown. (C) The ratio of CK18-M30/total CK18, an indicator of the fraction of apoptosis versus total epithelial cell death, is shown. (D) HMGB-1 levels in BALF from a COVID-19 mouse model. (E) Images and (F) densitometry of mixed lineage kinase domain-like (MLKL), p-MLKL, gasdermin D (GSDMD), and cleaved GSDMD immunoblots of the protein extracted from the lung of a COVID-19 mouse model are shown. (G) Representative images of the immunohistochemical analysis of p-MLKL and GSDMD in lung sections of mice are shown. Scale Bar = 50μm. Values are presented as means ± standard error. *p < 0.05, **p < 0.01, ***p < 0.0001.

Next, we determined whether PANoptosis (pyroptosis, apoptosis, and necroptosis)^39^, inflammatory programmed cell death pathways, are involved in alveolar epithelial cell death in the animal models of COVID-19. In the lung tissues of animal model of severe COVID-19, the levels of phospho-MLKL and cleaved GSDMD, executioners of necroptosis^40, 41^ and pyroptosis^42^, respectively, were significantly elevated compared with other two groups (Fig. 5E, F, Fig. S6). Additionally, cleaved caspase-3, an executioner of apoptosis, was also significantly increased in the COVID-19 animal models (Fig. S7). Immunohistochemical analysis demonstrated that both phospho-MLKL and GSDMD are localized within alveolar walls (Fig. 5G). Collectively, these results indicate that PANoptosis, including necroptosis and pyroptosis, contributes to alveolar epithelial cell death in COVID-19-induced ARDS.

### Anti-HMGB-1 antibody treatment attenuates alveolar tissue injury in animal models of severe COVID-19

Necrosis of alveolar epithelial cells seems to occur in very early stages of the pathogenesis of COVID-19-induced ARDS, and it is difficult to prevent alveolar epithelial necrosis prior to hospital admission. Therefore, we assessed whether inhibition of one of DAMPs, HMGB-1, attenuated alveolar tissue injury in a severe COVID-19 animal model. Treatment with the anti-HMGB-1 neutralizing antibody 4 h after intratracheal instillation of poly (I:C) and the SARS-CoV-2 spike protein significantly decreased BALF levels of leukocyte infiltration, total protein, ANG-2, total CK18 and CK-18 M30 (Fig. 6 A–F). On the other hand, the CK18-M30/total CK18 ratio was not affected by the anti-HMGB-1 treatment (Fig. 6G). These results suggest that DAMPs such as HMGB-1 are promising therapeutic targets that may be used to prevent the aggravation of COVID-19-induced ARDS after hospital admission.

**Figure 6.**
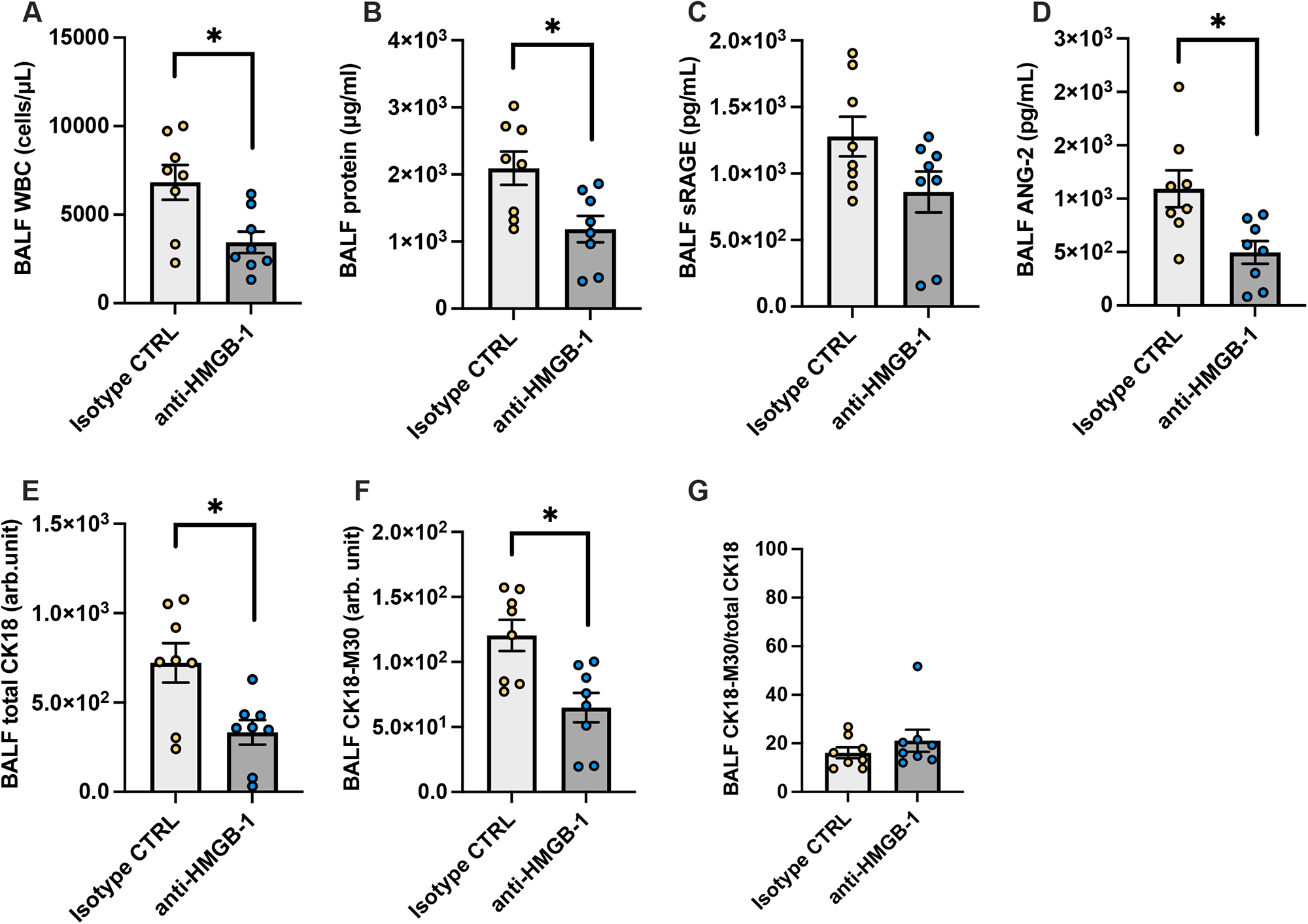
An analysis of effects of high mobility group box (HMGB)-1 neutralization on alveolar tissue injury in a mouse model of severe COVID-19. (A) White blood cell count, (B) total protein, (C) soluble receptors for advanced glycation end products (sRAGE), (D) angiopoietin (ANG)-2, (E) total CK18, (F) CK18-M30 levels, and (G) the ratio of CK18-M30/total CK18in BALF from the mouse model of severe COVID-19 treated with an anti-HMGB-1 neutralizing antibody or an isotype control antibody are shown. Values are presented as means ± standard error. *p < 0.05, **p < 0.01, ***p < 0.0001.

## Discussion

In the present study, we demonstrated that necrosis is the predominant form of alveolar epithelial cell death in COVID-19-induced ARDS. Moreover, two forms of programmed necrosis, necroptosis, and pyroptosis, were involved in the alveolar epithelial necrosis in mouse models of severe COVID-19. Animal experiments also suggested that DAMPs released from necrotic cells such as HMGB-1 are potential drivers of disease exacerbation in COVID-19-induced ARDS.

Alveolar tissue injury in severe COVID-19 is aggravated after passing the peak viral load^3, 4^. Therefore, the hyperinflammatory responses only against SARS-CoV-2 *per se* cannot fully explain the mechanisms underlying disease progression. Previous studies have reported that SARS-CoV-2-infected macrophages and monocytes undergo inflammasome activation and pyroptosis, potentially resulting in DAMPs release and excessive inflammation^43–45^. However, the pathological contribution of damages to non-immune cells remains nebulous. Recently, we have suggested that alveolar epithelial injury at a very early disease stage may trigger subsequent COVID-19 progression^5^. Herein, we show that the initial alveolar epithelial necrosis and subsequent release of DAMPs may also be a cause of excessive inflammation, which can progress even after viral loads have peaked. Not only cellular infection of SARS-CoV-2, but also inflammatory mediators, including TNF-α and IFN-γ, can cause programmed necrosis^39^. It is possible that programmed necrosis of immune and alveolar epithelial cells synergistically augment inflammatory responses and cell death or each other.

Both necrosis and apoptosis are involved in alveolar epithelial cell injury in ARDS^9^. Since apoptosis can be easily assessed using TUNEL staining or caspase detection, the contribution of alveolar epithelial apoptosis in ARDS has been extensively studied^46^. However, we have previously demonstrated that necrosis is the dominant form of alveolar epithelial cell death in LPS-induced ARDS by quantification of CK18-M30 and total CK18, which is equivalent to CK18-M65, in addition to cell labeling techniques^10^. In particular, quantification of CK18-M30 and M65 levels, using commercially available ELISA kit, can be applied for evaluation of epithelial apoptosis and necrosis in clinical setting. In fact, the patterns of epithelial cell death such as sepsis^47, 48^ and graft rejection after lung transplantation^49^ have been previously analyzed. To the best of our knowledge, this is the first study to suggest that necrosis is the predominant form of alveolar epithelial cell death in human ARDS. Further studies are warranted to identify alveolar patterns of epithelial cell death in ARDS that is induced by disease etiologies other than COVID-19.

Necrosis has been previously thought to cause accidental cell death; however, some forms of necrosis, referred to as programmed necrosis, are regulated via molecular pathways^7^. Several animal studies have demonstrated that programmed necrosis is involved in alveolar epithelial cell death in ARDS^50–53^. Moreover, studies have suggested that SARS-CoV-2 activates intracellular necroptosis and pyroptosis pathways^39, 44, 45, 54–56^, and that the circulating level of receptor-interacting protein kinase 3, a kinase required for necroptosis, is elevated in critically ill patients with COVID-19^57^. In line with findings of these studies, our animal experiments suggest that necroptosis and pyroptosis are involved in alveolar epithelial cell death in COVID-19 ARDS. Programmed necrosis is a response to eliminate SARS-CoV-2 infection, however, it can also cause excessive inflammation and subsequent tissue damages.

The release of DAMPs to extracellular spaces is a characteristic of necrosis that distinguishes it from apoptosis^58^. Several previous studies have also reported that circulating levels of DAMPs, such as HMGB-1^59–62^, histone^62, 63^, cell free-DNA^62, 63^, mitochondrial DNA^64–66^ and S100 proteins^67, 68^ are elevated in severe COVID-19. Alveolar epithelial necrosis likely occurs very early in COVID-19 progression^5, 6^ and potentially contributes to disease progression. Moreover, it is possible that these DAMPs can be released from necrosis of immune cells such as macrophages^44, 45^. In both cases, the prevention of necrosis prior to the appearance of clinical symptoms is difficult; therefore, a strategy for preventing DAMPs-mediated disease aggravation is needed. HMGB-1 is the most extensively studied DAMPs which drives tissue injury during ARDS or sepsis; results from our animal study demonstrated that the inhibition of HMGB-1 efficiently attenuates disease progression. However, it remains unclear as to the type of DAMPs with the most contribution to disease progression. Further studies, including clinical trials, are warranted to investigate the clinical efficacy of DAMP inhibition in patients with COVID-19 ARDS.

In the present study, an animal model mimicking COVID-19 was established by administering the SARS-CoV-2 spike protein combined with poly (I:C), similar to previous studies^18, 69, 70^. The animal models treated with infectious strains of SARS-CoV-2 are often ideal models for COVID-19; however, using infectious viruses in animal experiments can be difficult. First, SARS-CoV-2 infection does not occur in wild-type mice or rats. For infection to occur, expression of the human ACE receptor is needed. Second, appropriate facilities and equipment are needed for meet all the safety requirement when working with an infectious agent. Although the pathogenicity of SARS-CoV-2 is complex, stimulation of pathogen-associated pattern recognition receptors including toll-like receptors^26, 27^ and retinoic acid-inducible gene-I receptors^28^ by viral components is the principal driver of lung inflammation, and subsequent alveolar tissue damage. In the present study, the bioinformatic analysis of lung tissue transcriptomes demonstrated that the COVID-19 mimicking animal model with SARS-CoV-2 spike protein and poly (I:C) recapitulated key biological responses in inflammatory and cell death pathways induced by SARS-CoV-2^32–37^. Moreover, alveolar cell death patterns in our COVID-19 animal model were similar to those observed in human COVID-19^29, 30^. Our results highlight the utility of investigating the pathophysiology and treatment of COVID-19 using animal models established with components of SARS-CoV-2.

Our data suggest that plasma M30/M65 ratio (an indicator of apoptosis in relation to total levels of epithelial cell death) is a potential marker of COVID-19 severity. Our findings agreed with a previous study that showed M30/M65 ratios of hospitalized COVID-19 patients were lower than those of non-hospitalized patients^71^. Additionally, different subtypes of COVID-19 respond differently to treatments ^72, 73^. The M30/M65 ratio may serve as a marker for selecting patients likely to benefit from anti-DAMPs treatment.

In summary, our data indicate that necrosis, including necroptosis and pyroptosis, is the predominant form of alveolar epithelial cell death in COVID-19-induced ARDS. The DAMPs released from necrotic alveolar epithelial cells are potential drivers of progressive alveolar tissue damage in COVID-19, and hence are promising targets for preventing the aggravation of ARDS in patients with COVID-19.

## Limitations of the study

This study has some limitations. First, only patients admitted to a single center were included in the analysis due to the limited availability of clinical samples. Further studies with samples from multiple centers in different countries are warranted. Second, serum samples were used in the most parts of the human study, as the bronchoalveolar lavage fluids were available only from limited number of severe COVID-19 patients. However, severe organ injury was almost limited to lungs in the present cohort, tissue injury markers in serum samples might well reflect the pulmonary pathology. Third, our animal model was created by exposing mice to components of SARS-CoV-2, and not an infectious strain of SARS-CoV-2. Despite this, the observations of a COVID-19-like pathology by this study and previous reports ^18, 70^ support the use of the animal model.

Importantly, use of a non-infectious model is convenient for laboratories that do not specialize in infectious disease research. Fourth, the efficacy of inhibiting only a single DAMP, HMGB-1, was evaluated. Several types of DAMPs are released from necrotic cells; therefore, whether the what types of DAMPs are the primary therapeutic targets for COVID-19 remain to be determined.

## Data Availability

All data produced in the present study are available upon reasonable request to the authors.

## Acknowledgements

We would like to thank Ms. Yuki Yuba and Ms. Akiko Adachi (Department of Anesthesiology and Critical Care Medicine, Yokohama City University) for technical assistance. We are deeply grateful to Dr. Jordan A. Ramilowski and Dr. Tomohiko Tamura (Advanced Medical Research Center, Yokohama City University) for instructing bioinformatic analysis through MEXT Joint Usage/Research Center Program at the Advanced Medical Research Center, Yokohama City University. We also thank the Department of Emergency Medicine, Department of Microbiology, and Yokohama City University Center for Novel and Exploratory Clinical Trials for collecting and providing the blood and BALF samples. This work was partly supported by AMED under Grant Number JP20he0522001, Yokohama Foundation for Advancement of Medical Science under Research Grant for COVID-19, JSPS KAKENHI Grant Number 21K16575 and 22K09146, and Grant for 2020-2021 Research Development Fund of Yokohama City University.

## Authors’ contributions

Conceptualization and study design: KT. Acquisition of clinical study data: KT, NY, MA. Supervision of clinical sample collection: MN, IT. Acquisition of animal experiments data: KT, NT. Bioinformatic analysis: KT. Analysis and interpretation of data: KT, NY, NT, TM, TG. Manuscript Writing: KT. Manuscript Editing: All investigators.

## Methods

### Resource Availability

#### Lead contact

Further information and requests for resources and reagents should be directed to and will be fulfilled by the lead contact, Kentaro Tojo.

#### Materials availability

This study did not generate new unique reagents.

#### Data and code availability

The RNA-seq data have been deposited at GEO and are publicly available as of the date of publication. Accession numbers are listed in the key resources table. This paper analyzes existing, publicly available data. These accession numbers for the datasets are listed in the key resources table. Any other data reported in this paper will be shared by the lead contact upon request. This paper does not report original code. Any additional information required to reanalyze the data reported in this paper is available from the lead contact upon request.

### Experimental Model and Subject Details

#### Human subjects

Forty-eight adult patients with COVID-19 (male = 36; female = 12; median age = 68.5) who were admitted to Yokohama City University Hospital from January 2020 to January 2021 and 18 healthy controls (male = 12; female = 6; median age = 52.0) were included in the serum analysis. Six patients with COVID-19-induced ARDS (male = 5; female = 1; median age =58.0) who were admitted to the Yokohama City University Hospital from April 2021 to January 2022 were included in the bronchoalveolar lavage fluid analysis. The study protocol was reviewed and approved by the institutional review board of Yokohama City University Hospital (B200700100, B200200048). The requirement for informed consent was waived due to the observational nature of the study.

#### Animals

Male specific-pathogen-free C57BL/6J mice aged 8–10 weeks that were purchased from Japan SLC (Shizuoka, Japan) and housed under a 12-h light/dark cycle with food and water available *ad libitum*. All animal experimental protocols were approved by the Animal Research Committee of the Yokohama City University. All experiments were performed in accordance with the relevant regulatory standards.

### Method Details

#### Clinical study design

In this single-center, retrospective, prospective observational study, we analyzed serum samples of adult patients with COVID-19 who were admitted to Yokohama City University Hospital from January 2020 to January 2021 and healthy controls matched as closely as possible for age and sex. Inclusion criteria for COVID-19 patients were, as follows: 1) a diagnosis of COVID-19 based on a positive real-time polymerase chain reaction test, 2) age ≥ 18 years, and 3) available residual serum samples. Additionally, we analyzed BALF samples from patients with COVID-19-induced ARDS who were admitted to the Yokohama City University Hospital from April 2021 to January 2022. ARDS was diagnosed based on the Berlin definition. The study protocol was reviewed and approved by the institutional review board of Yokohama City University Hospital (B200700100, B200200048). The requirement for informed consent was waived due to the observational nature of the study. Some preliminary data from retrospectively collected samples have been previously published^5^.

#### Clinical data collection

The following clinical data measured during the first 8 days of hospital admission were retrospectively collected from the medical charts of included patients: basal characteristics, vital signs, laboratory tests, and blood gas analysis findings.

#### Human serum sample analysis

Residual serum samples collected from patients with COVID-19 after daily laboratory tests were frozen for future use. Concentrations of human serum soluble receptors for advanced glycation end products (sRAGE) (DY1145, R&D systems, Minneapolis, MN), angiopoietin (ANG)-2 (DY623, R&D Systems), surfactant protein (SP)-D (DY1920, R&D Systems), cytokeratin (CK)18-M65 (M65 ELISA, #10040, VLVBio AB, Nacka, Sweden), CK18-M30 (M30-Apoptosense ELISA Kit, #10011, VLVBio), and HMGB-1 (#381-10531, Fuso, Osaka, Japan) were measured using commercially available enzyme-linked immunosorbent assay (ELISA) kits according to the manufacturer’s instructions. The ratio of CK18-M30/M65 was calculated, and when the value exceeded 100%, it was regarded as 100%.

Th initial concentrations of these markers in ARDS and non-ARDS patients at admission (on the first or second hospital day) and healthy controls were compared. Further, temporal changes in levels of the markers were assessed in patients with ARDS throughout the 8-day period following hospital admission. In cases in which values were determined twice per day, mean values were used. In cases in which only a single value was available, the value was used.

#### Analysis of BALF samples from COVID-19-induced ARDS patients

BALF samples were obtained from six COVID-19-induced ARDS patients. A fiberoptic bronchoscope was wedged in a lateral or medial segmental bronchus of the right middle lobe, and lavage was performed using three aliquots of 50 mL of sterile isotonic sodium chloride solution. The collected BALF was centrifuged at 300 × *g* for 5 min at 4 : and the supernatant was stored at -80 c: until analyses. The levels of CK18-M65 and CK18-M30 were quantified using ELISA, while the ratio of CK18-M30/M65 was calculated as described above.

#### Animal experiments

All animal experimental protocols were approved by the Animal Research Committee of the Yokohama City University. Male specific-pathogen-free C57BL/6J mice aged 8–10 weeks that were purchased from Japan SLC (Shizuoka, Japan) were used for all animal experiments. Mice were housed under a 12-h light/dark cycle with food and water available ad libitum.

A mouse model mimicking COVID-19 was established based on previous reports.^18, 69^ Intratracheal administration of polyinosinic:polycytidylic acid (poly (I:C)) (P1530, Sigma-Aldrich, St. Louis, MO, USA) and the SARS-CoV-2 spike protein (Z03481, Lot B2103045, GenScript, Piscataway, NJ) was performed via the exposed trachea through a small incision at the front of the neck. During the procedure, mice were placed under general anesthesia using intraperitoneal ketamine and xylazine. The mice were euthanized 24 h after intratracheal instillation, and lung tissues and bronchoalveolar lavage fluid (BALF) samples were collected as previously described^10, 74^.

In a preliminary experiment, nine mice were randomly allocated into the following three groups (n = 3 per group): control, Poly (I:C), and Poly (I:C) combined with the SARS-CoV-2 spike protein. The poly (I:C) group received 250 μ intratracheal poly (I:C) dissolved in 100 μ phosphate buffered saline (PBS), and poly (I:C) combined with SARS-CoV-2 group received 50 μg SARS-CoV-2 spike protein with 250 μg poly (I:C) dissolved in 100 μ PBS. The control group received 100 μL PBS intratracheally. Based on the results of preliminary experiments, mild and severe lung injuries mimicking COVID-19 were evaluated. Twelve mice were randomly allocated into the following three groups (n = 4 per group): control, mild COVID-19, and severe COVID-19. The severe COVID-19 group received 50 μg SARS-CoV-2 spike protein with 250 μg poly (I:C) dissolved in 100 μL PBS, whereas the mild COVID-19 group received 10 μg SARS-CoV-2 spike protein and 50 μg Poly (I:C) in 100 μL of PBS. The control group received 100 μL PBS intratracheally.

Finally, we evaluated effects of anti-HMGB-1 neutralizing antibodies on a severe COVID-19 animal model. Six mice were randomly allocated to anti-HMGB-1 antibody or isotype control groups (n = 8 per group). The severe COVID-19 animal model was established as described above. Then, 4 h after intratracheal instillation, 100 μg anti-HMGB-1 neutralizing (ARG66714, Arigo Biolaboratories, Hsinchu City, Taiwan) or isotype control antibodies dissolved in 100 μL PBS were intravenously administered via the tail vein under isoflurane anesthesia.

#### Analysis of the BALF of mouse models of COVID-19

Leukocytes from the BALF of mice were stained with Samson’s solution and counted. Protein concentrations in BALF were quantified using a bicinchoninic acid assay. Concentrations of sRAGE (DY1179, R&D systems), ANG-2 (MANG20, R&D systems), CK18-M30 (CSB-E14265M, CUSABIO, Houston, TX), total CK18 (CSB-E17158M, CUSABIO), and HMGB-1 (ARG81310, Arigo Biolaboratories) were measured using ELISA kits in accordance with the manufacturer’s instructions. Cytokines and chemokines were comprehensively analyzed using semiquantitative multiplex cytokine assay kits (ARY006, R&D systems) in accordance with the manufacturer’s instructions.

#### Western blotting

The total proteins from mouse lung tissues were extracted using trichloroacetic acid-acetone, while the extracted proteins were solubilized and quantified using a bicinchoninic acid assay. A certain amount of protein (MLKL: 5 μg, GSDMD and caspase-3: 2 μg, cleaved GSDMD: 20 μg, pMLKL: 30 μg, and cleaved caspase-3: 50 μg of proteins) was separated by sodium dodecyl sulphate-polyacrylamide gel electrophoresis and transferred to polyvinylidene fluoride (PVDF) membranes. Proteins were detected using primary antibodies against mixed lineage kinase domain-like (MLKL) (#28640, Cell Signaling Technology, Danvers, MA, dilution: 1:2,000), pospho-MLKL (#37333, Cell Signaling Technology, 1:1,000), gasdermin D (GSDMD) (ab219800, Abcam, Cambridge, UK, 1:3,000), cleaved n-terminal GSDMD (A20197, ABclonal, Woburn, MA, 1:1,000), caspase-3 (#9662 Cell Signaling Technology, 1:1,000), cleaved caspase-3 (#9661, Cell Signaling Technology, 1:1,000) and horseradish peroxidase-conjugated secondary goat anti-rabbit IgG antibodies (170-6515; Bio-Rad, Hercules, CA)^74^. Equality of protein loading was confirmed by total protein staining (in case of protein amounts >10 μg) (Reversible Protein Stain Kit for PVDF Membranes, 24585, Thermo Fisher Scientific, Waltham, MA) or beta-actin (A5411, Sigma-Aldrich, 1:10,000) staining (in case of protein amounts < 10 μg). The density of each protein was determined using ImageJ software (National Institutes of Health, Bethesda, MD).

#### Histological analysis

Lung tissues of mice were fixed using 4% paraformaldehyde at 20 cm H_2_O pressure and embedded in paraffin for histopathological examination^74^. Lung tissue sections were stained with hematoxylin and eosin for tissue injury evaluation. Lung tissue sections were stained with anti-pMLKL (ET1705-51, HUABIO, Woburn, MA, dilution: 1:100) and anti-GSDMD n-terminal (ER1901-37, HUABIO, 1:200) antibodies according to the manufacturer’s instructions.

#### RNA-seq of lung tissues

The total RNA was extracted from lung tissues of severe COVID-19 model and control mice using spin column (FastGene RNA, Nippon Genetics, Tokyo, Japan). The RNA-seq library was prepared using TruSeq stranded mRNA (Illumina, San Diego, CA) and was sequenced with NovaSeq 6000 (Illumina) by Macrogen Japan Corp. (Tokyo, Japan).

#### Publicly available RNA-seq data

We searched the National Center for Biotechnology Information Gene Expression Omnibus database using the following keywords: ((SARS-CoV-2) AND (mice OR mouse) AND (lung OR lungs)) AND “Mus musculus”[porgn: txid10090] and found 6 RNA-seq datasets analyzing early phase (day 1-5 post infection) lung transcriptomes of mice infected with SARS-CoV-2. The details of the includes studies are provided in Table S2. We compared lung tissue transcriptome data between days 1–5 post-infection animals and the control among each dataset. In GSE154104 dataset, data of animals with days 2 and 4 post-infection were combined and compared with the control.

#### Bioinformatic analysis of lung transcriptomes

The obtained FASTQ files of RNA-seq data including our own data and publicly available data were quality checked and trimmed with TrimGalore (Version 0.6.7, Babraham Bioinformatics). Thereafter, transcript quantification was performed with Salmon (Version 1.9.0)^75^, and gene-level read counts were obtained with tximport R package (Version 1.24.0)^76^ in R version 4.2.0 (The R Foundation for Statistical Computing). Comparison of gene expressions between COVID-19 model and the control in each dataset was performed with DESeq2 R package (Version 1.36.0)^77^. The gene ranking in each dataset was determined based on p-value and gene set enrichment analysis (GSEA) performed using fGSEA R package (Version 1.22.0)^78^.

In reciprocal GSEA, an upregulated or downregulated gene set in each data set was defined as top 100 upregulated or downregulated genes. The enrichment of each gene set within the pre-ranked gene lists of the other data sets was evaluated. GSEA for REACTOME pathway gene sets was also performed, and normalized enrichment scores (NESs) in each dataset were obtained. We selected pathways significantly enriched at least in one dataset, and the correlation between NESs in our own dataset and mean NESs in other 6 datasets of the infection models was analyzed. Moreover, log2 fold changes of the genes involved in the key inflammatory and cell death pathways in REACTOME database were compared between the model in the present study and the infection models.

### Quantification and Statistical Analysis

Statistical analyses were performed using Prism 9 software (GraphPad, La Jolla, CA). Values of P < 0.05 were considered statistically significant. Data from clinical studies were presented as medians with interquartile ranges (IQRs) and analyzed as non-parametric data. Comparisons between basal characteristics and laboratory and physiological values among patients with and without ARDS were performed using Mann−Whitney or Fisher’s exact tests. Comparisons of the serum levels of tissue injury and cell death markers among patients with and without ARDS, and healthy controls were performed using Kruskal−Wallis analysis followed by Dunn’s multiple comparison test. Temporal changes in alveolar tissue injury markers were assessed using the Friedman test, and days in which alveolar tissue injury markers peaked were compared using Kruskal−Wallis analysis followed by Dunn’s multiple comparison test. The data obtained from animal experiments were log-transformed and presented as means ± standard deviation and analyzed using the Student’s *t*-test or one-way analysis of variance followed by Tukey’s multiple comparison test. Multiple cytokine and chemokine assays were analyzed using multiple *t*-tests via the false discovery rate approach comprised of the two-stage step-up method of Benjamini, Krieger, and Yekutieli^79^. The false discovery rate was set at 5%.

