## Supplementary Files for "Early alveolar epithelial cell necrosis is a potential driver of COVID-19-induced acute respiratory distress syndrome"

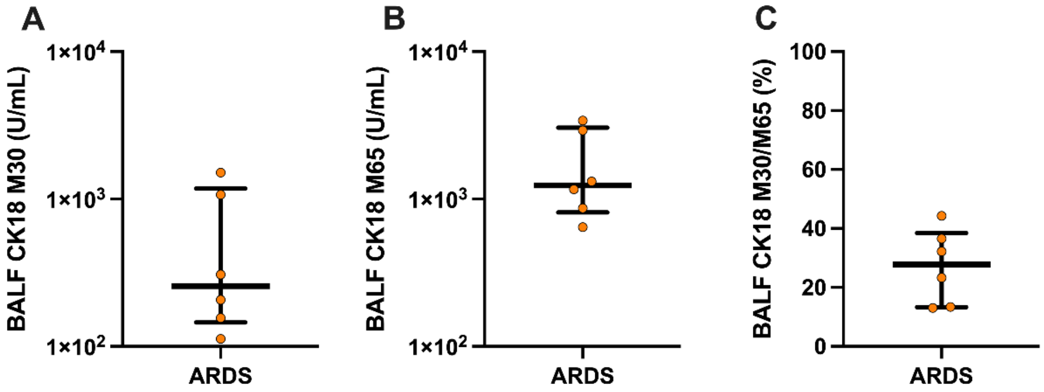


**Figure S1. Levels of epithelial cell death markers in BALF samples, Related to Figure 2.** Levels of (A) CK18-M30, an epithelial apoptosis marker; (B) CK18-M65, an epithelial total cell death marker; (C) CK18-M30/M65 ratio, an indicator of the fraction of epithelial cells undergoing apoptosis versus all types of cell death in BALF samples of COVID-19 patients with ARDS

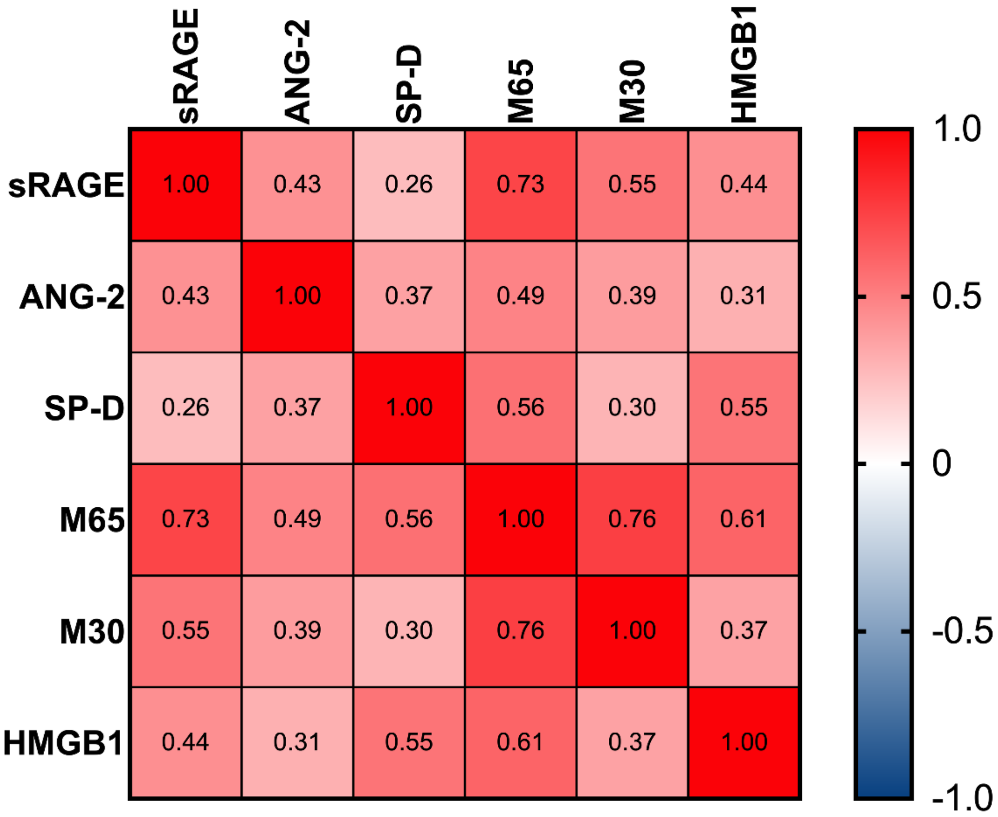


**Figure S2. Pearson correlation coefficients among circulating markers, Related to Figure 1 and 2.** The serum levels of each marker were log-transformed and then Pearson's correlation analyses were performed.

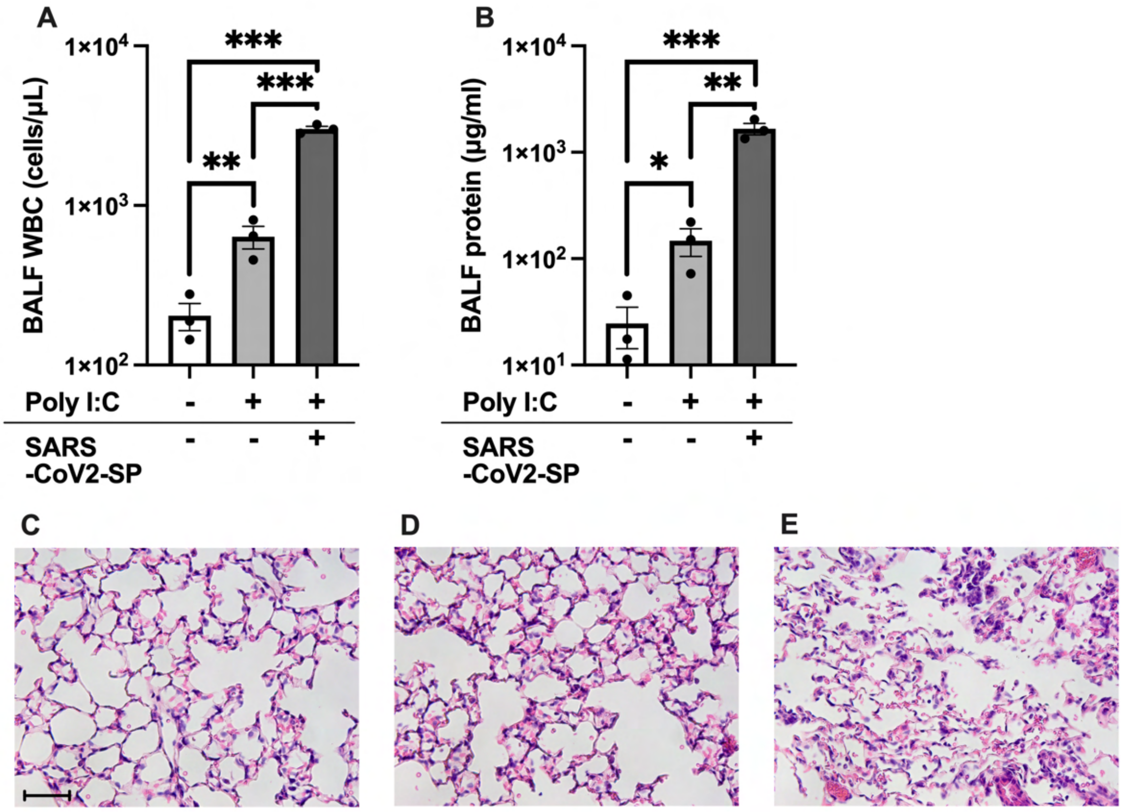


**Figure S3. Effects of intratracheal administration of the severe acute respiratory syndrome coronavirus 2 (SARS-CoV-2) spike protein combined with polyinosinic:polycytidylic acid (poly (I:C)), Related to Figure 3.** Levels of (A) white blood counts and (B) total proteins in the bronchoalveolar lavage fluid of mice intratracheally injected with phosphate buffered saline, poly (I:C) or SARS-CoV-2 spike proteins combined with poly (I:C) are shown. (C) Representative images of lung tissue sections stained with hematoxylin and eosin. Scale Bar = 50μm. The values are presented as means ± standard error. *p<0.05, **p<0.01, ***p<0.0001

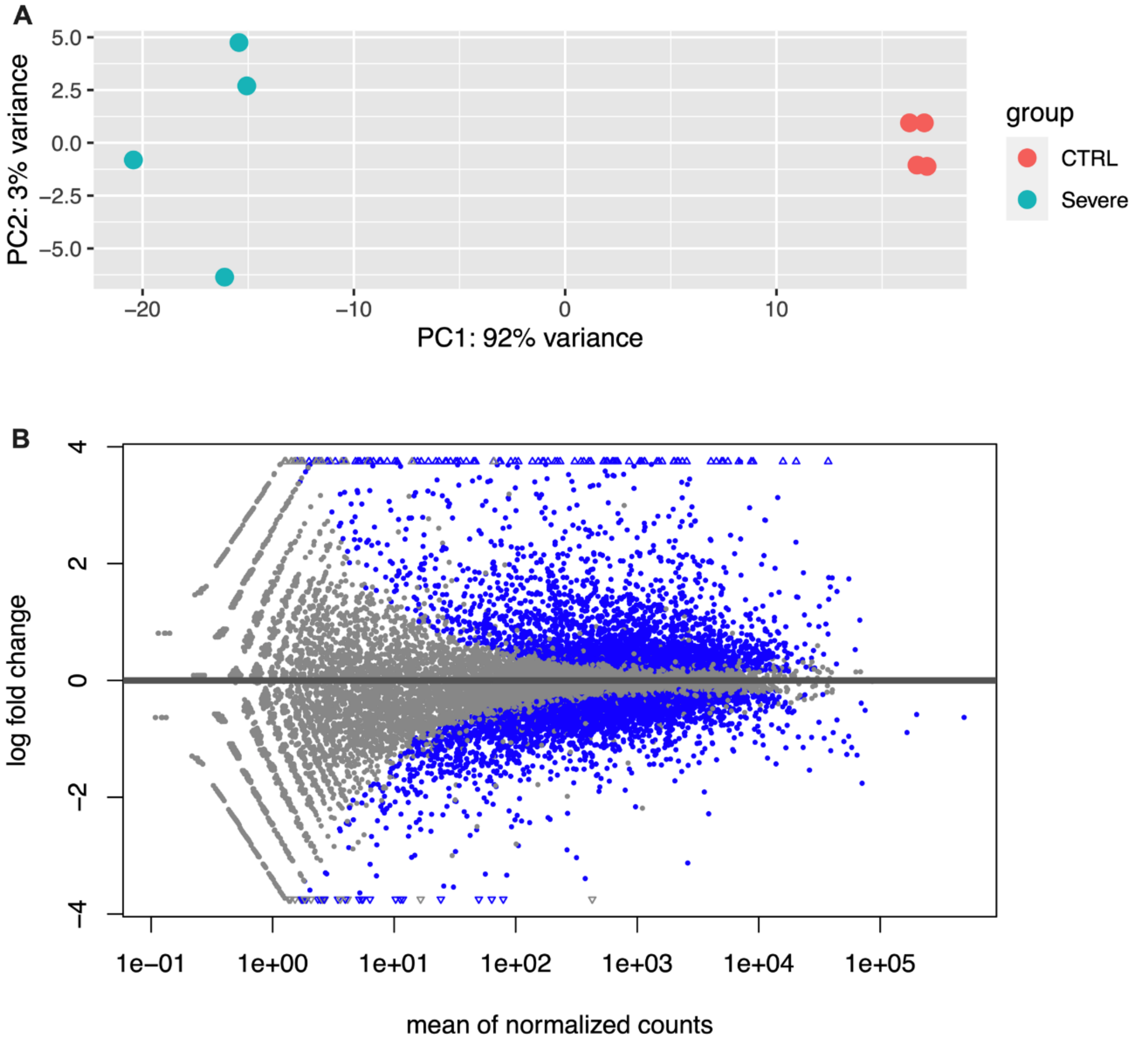


**Figure S4. Analysis of RNA-seq data of the COVID-19 model using SARS-CoV-2 spike protein and poly (I:C) in the present study, Related to Figure 4.** (A) Principal component analysis and (B) MA-plot comparing the severe COVID-19 model and the control

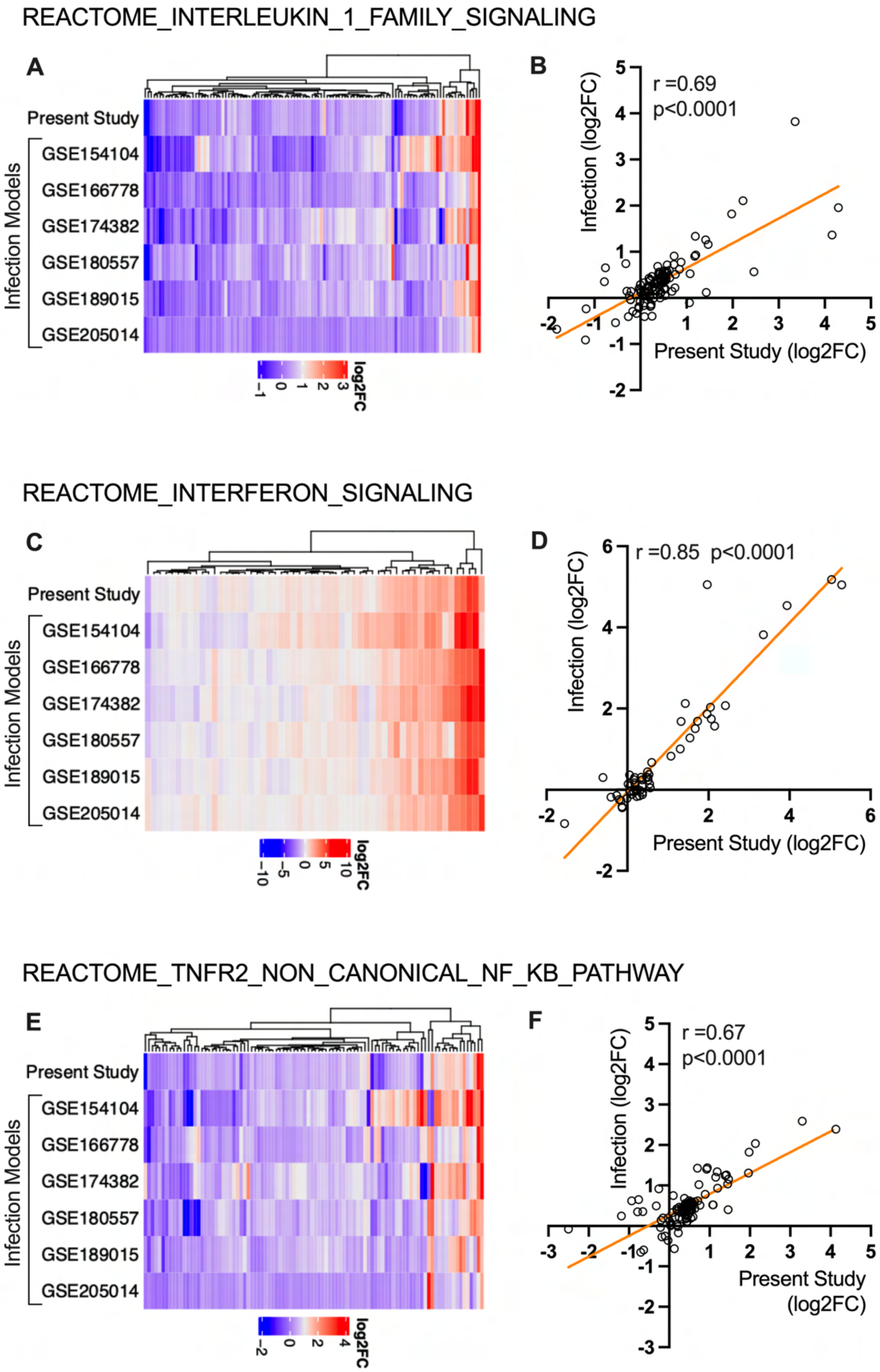


**Figure S5** (continued on next page, legend follows).

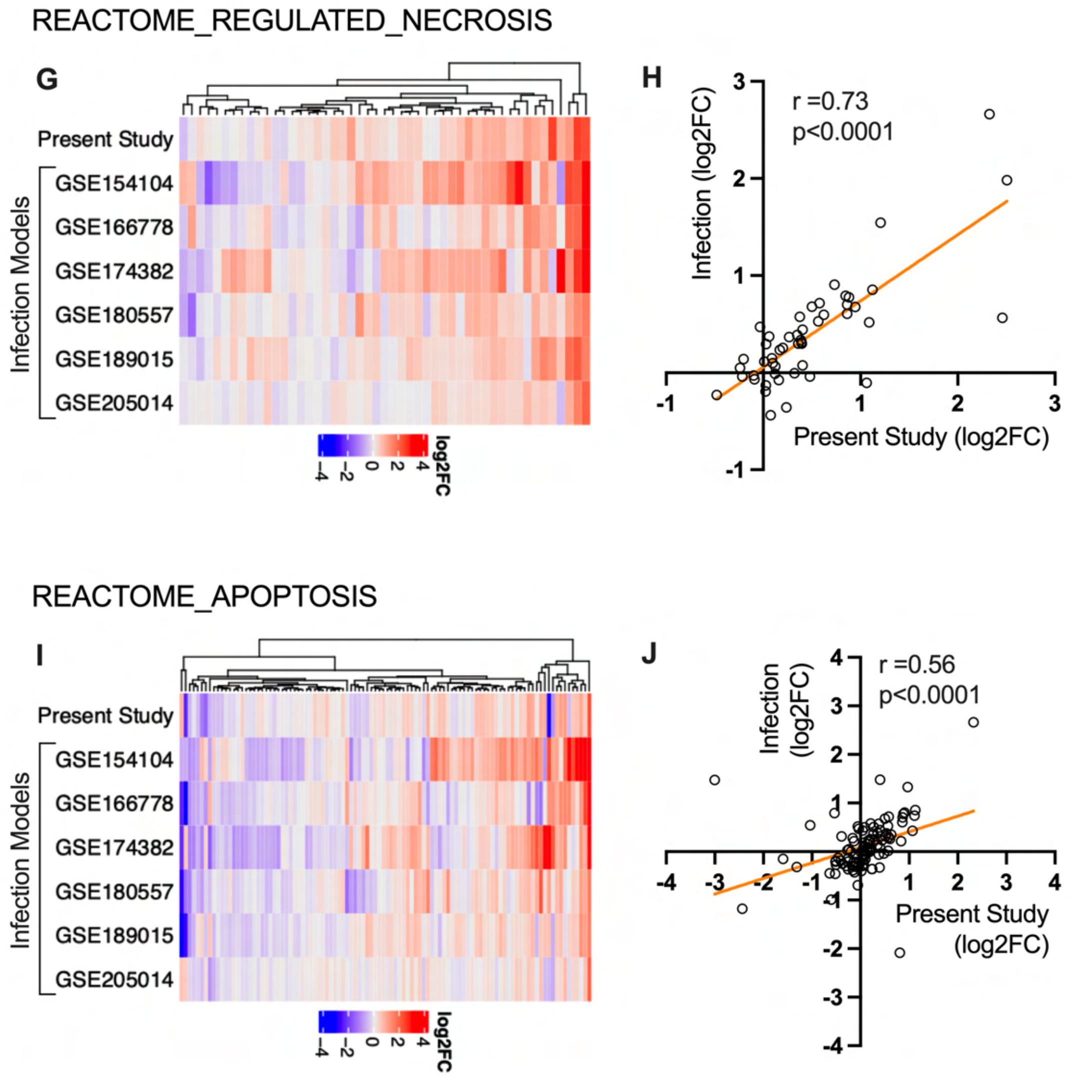


**Figure S5. Analysis of gene expression patterns in the Reactome, Related to Figure 4.** (A, B) Interleukin-1 family signaling, (C, D) interferon signaling, (E. F) TNFR2 non-canonical NFκB pathway, (G, H) regulated necrosis, and (I, J) apoptosis pathways. Heatmaps of gene expression patterns (A, C, E, G, and I) and correlation analysis between the $\log_2$ fold-changes in the present study model and mean $\log_2$ fold-changes in the infection models (B, D, F, H, and J) are shown

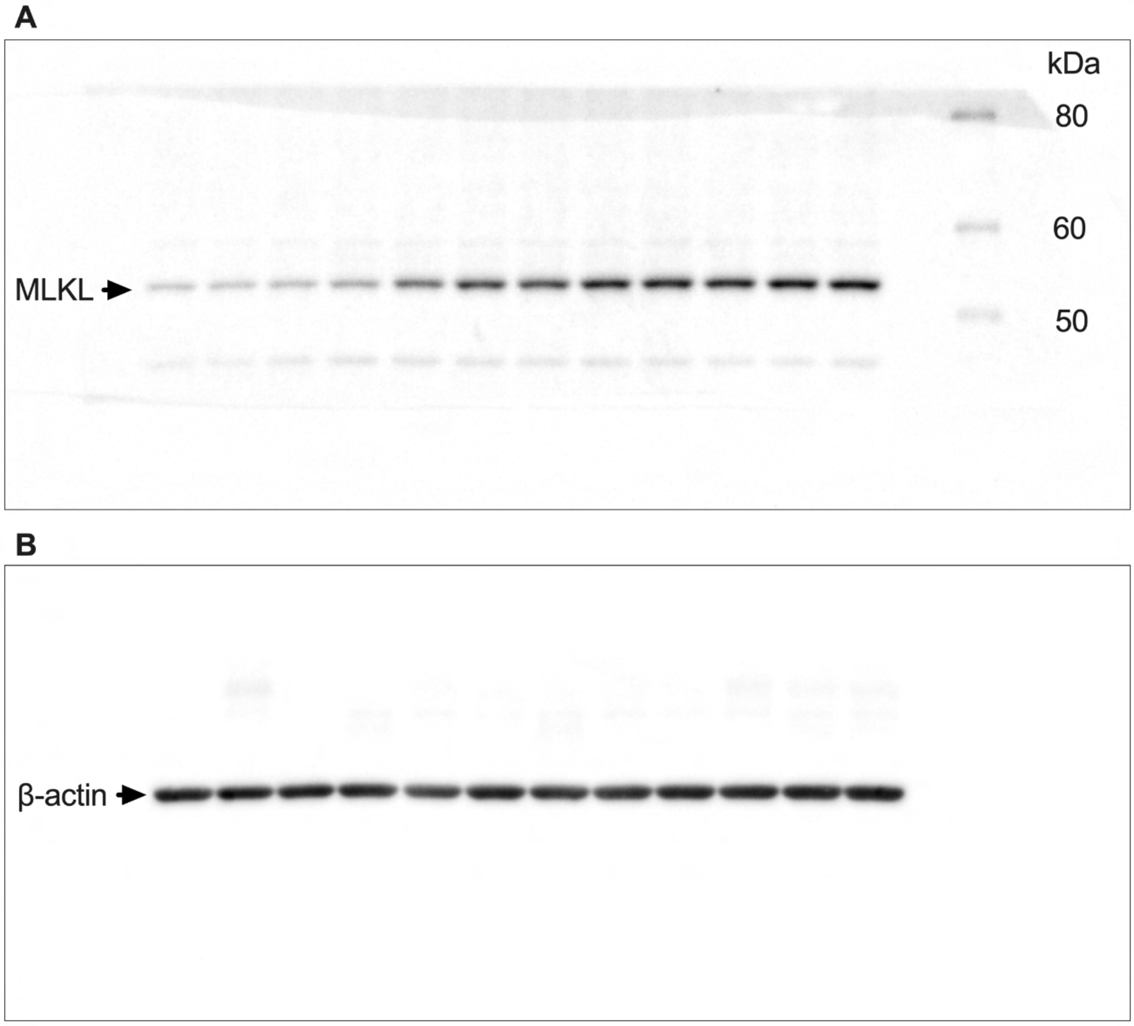


**Figure S6** (continued on next page, legend follows).

C

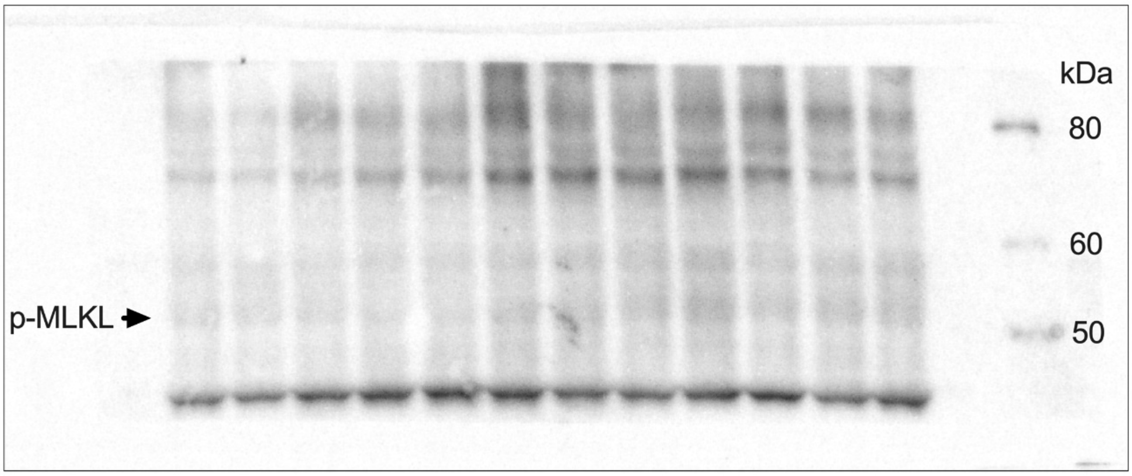


D

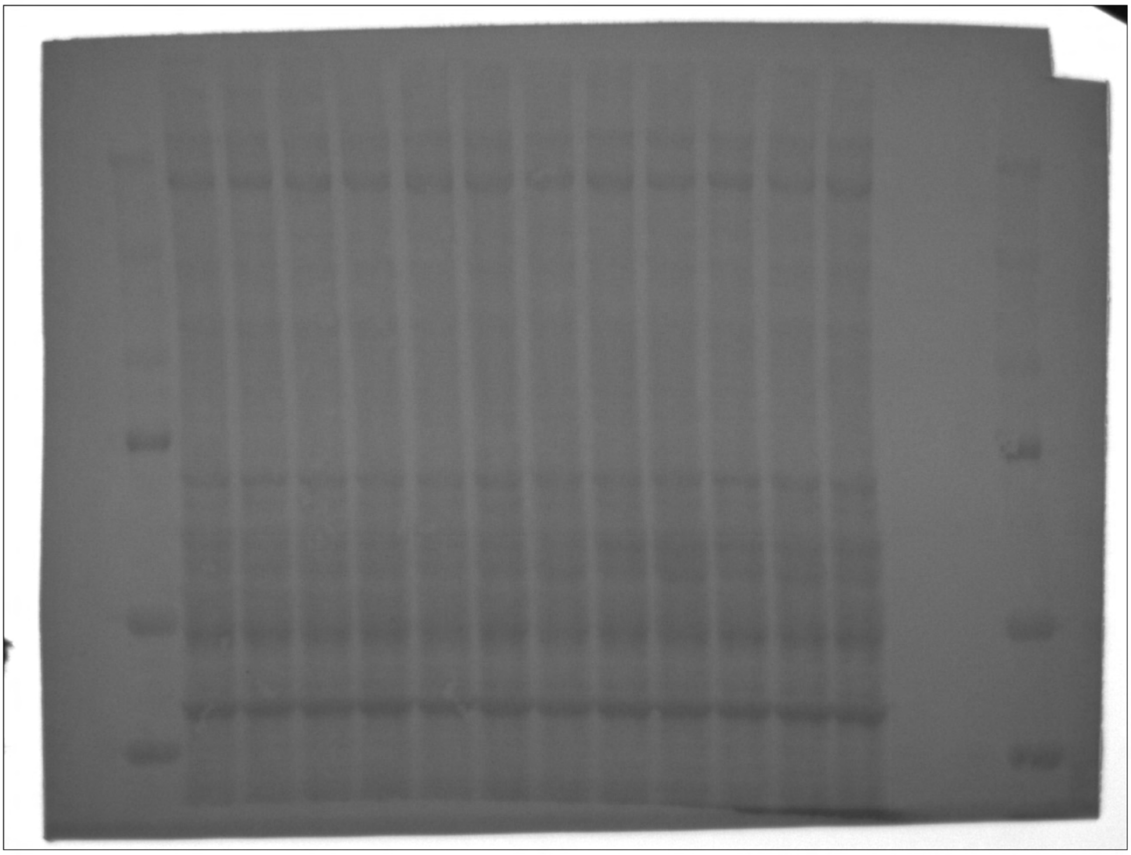

**Figure S6** (continued on next page, legend follows).

**E**

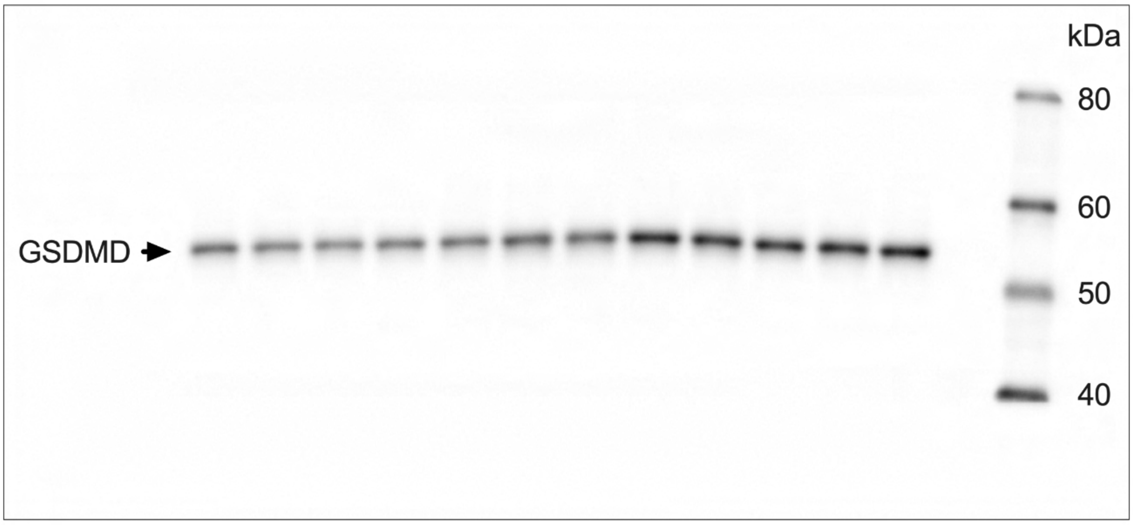


**F**

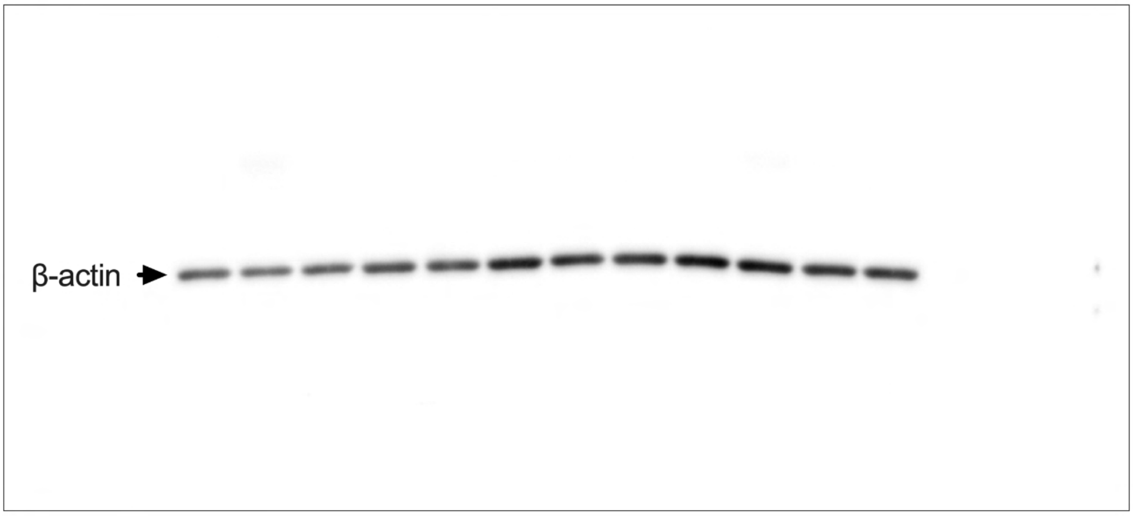


**Figure S6** (continued on next page, legend follows).

G

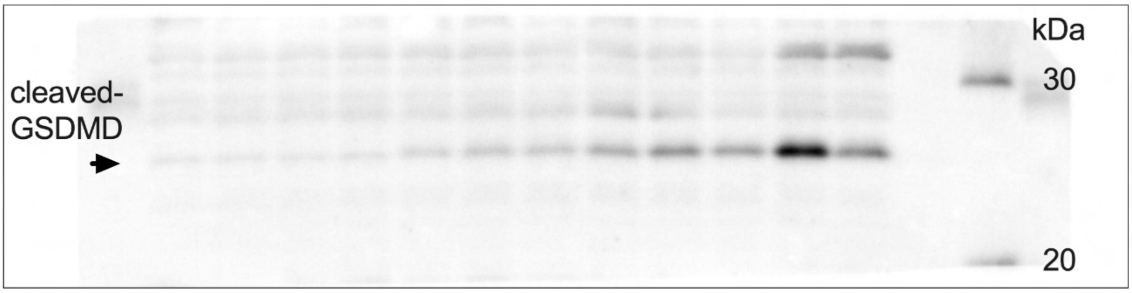


H

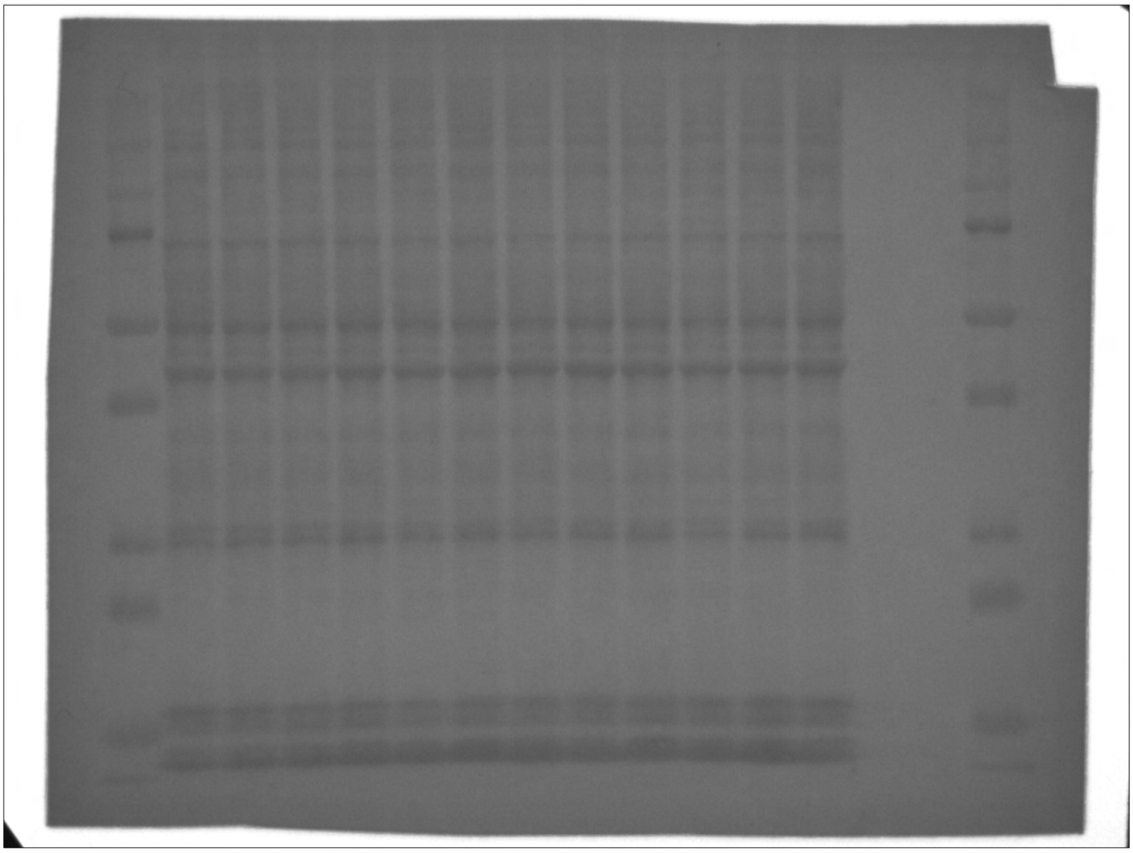

**Figure S6. Uncropped images of immunoblots of target proteins and loading controls, Related to Figure 5.** Equality of protein loading was confirmed by beta-actin staining (in case of protein amounts <10 µg) after quenching horseradish peroxidase using hydrogen peroxide or total protein staining (in case of protein amounts >10 µg). Membranes of (A, B) mixed lineage kinase domain-like (MLKL) and (C, D) phosphorylated MLKL, (E, F) gesdermin D (GSDMD), and (G, H) cleaved GSDMD are shown

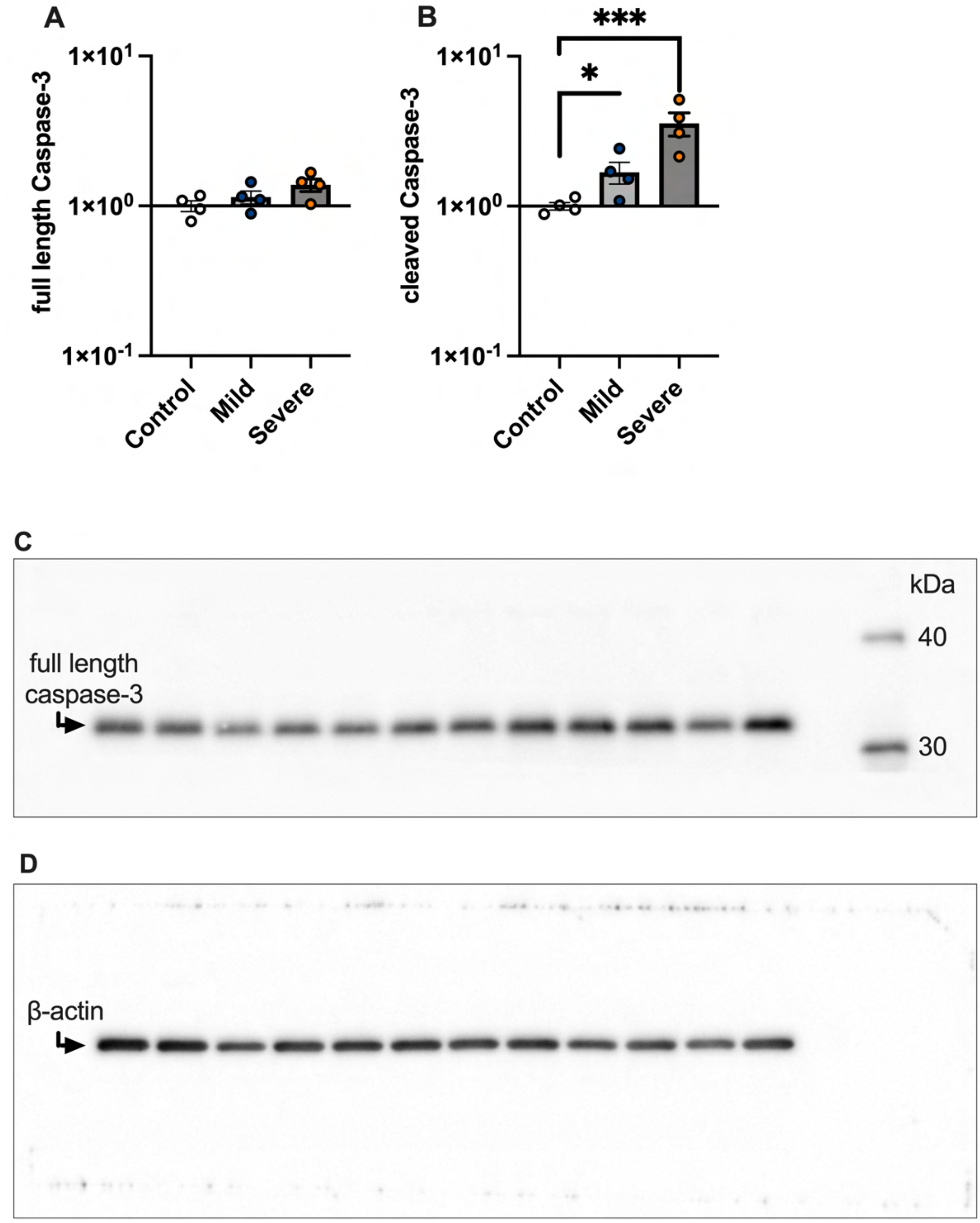


**Figure S7** (continued on next page, legend follows).

E

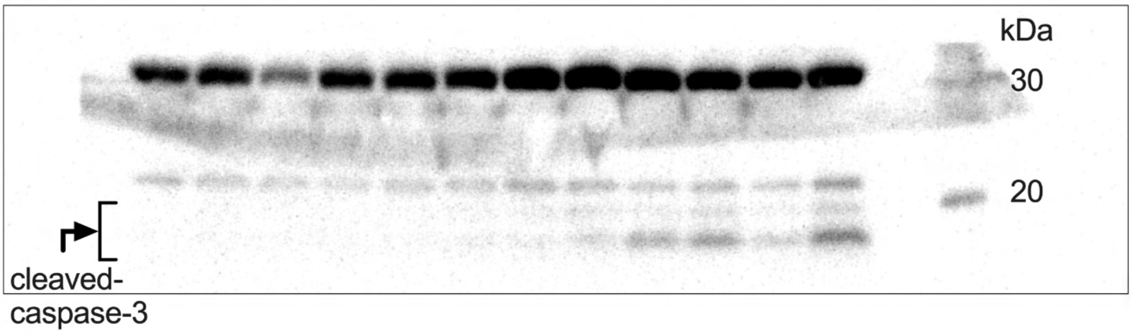


F

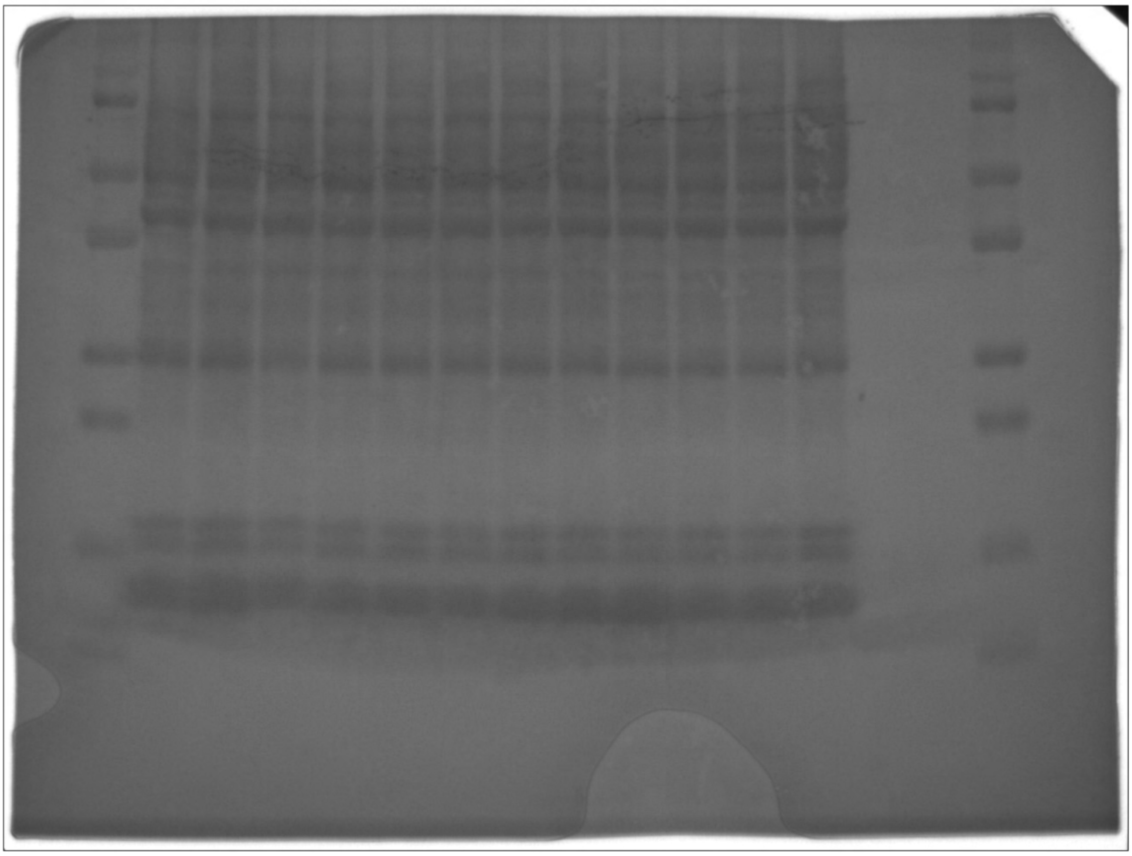

**Figure S7. Immunoblot analysis of full length and cleaved caspase-3, Related to Figure 5.** (A, B) Densitometry and (C-F) Uncropped images of full length and cleaved caspase-3 immunoblots of the protein extracted from the lung tissue of a COVID-19 mouse model. Equality of protein loading was confirmed by beta-actin staining (full length caspase-3) after quenching horseradish peroxidase using hydrogen peroxide or total protein staining (cleaved caspase-3).

| | |
|---|---|
| **Age (years)** | 58 (55-71) |
| **Males/Females** | 5/1 |
| **APACHE2 score** | 14.0(10.0-15.5) |
| **P/F ratio at admission** | 154.5 (132.0-215.2) |
| **Mechanical ventilation use** | 6 (100.0%) |
| **In hospital mortality** | 1 (16.7%) |
| **Laboratory data on admission** | |
| **WBC count (/μL)** | 8950 (3300-13700) |
| **Lymphocyte count (/μL)** | 563 (435-916) |
| **Platelet count (×$10^3$/ μL)** | 165.5 (110.3-195.0) |
| **D-dimer (μg/mL)** | 1.30(1.14-3.32) |
| **CRP (mg/dL)** | 4.69 (3.73-17.12) |
| **Creatinine (mg/dL)** | 0.81 (0.57-4.23) |
| **Total bilirubin (mg/dL)** | 0.40 (0.28-0.45) |

**Table S1. Clinical characteristics in ARDS patients (n=6) with COVID-19 whose BALF samples were collected, Related to Figure 2 and Figure S1.** Data are presented as count (%) or median (IQR).

| Accession Number | Strain | Age | Sex | Disease Tissue | Control Tissue |
|---|---|---|---|---|---|
| GSE216644 (The Present Study) | C57BL/6J | 8 weeks | Male | Instillation of SARS-CoV-2 spike protein and poly(I:C) | Instillation of Phosphate-buffered saline |
| GSE154104 | K18-hACE2 | 8 weeks | Male/ Female | SARS-CoV-2-infected lung | Non-infected lung |
| GSE166778 | BALB/c | 6 weeks | Male | Mouse-adapted SARS-CoV-2-infected lung | Non-infected lung |
| GSE174382 | K18-hACE2 | 8-10 weeks | Male | SARS-CoV-2-infected lung | Non-infected Lung |
| GSE180557 | K18-hACE2 | 8-16 weeks | Male | SARS-CoV-2-infected lung | Non-infected Lung |
| GSE189015 | CD-1 | 8-10 weeks | Female | SARS-CoV-2-infected lung | Non-infected Lung |
| GSE205014 | K18-hACE2 | 9 weeks | Female | SARS-CoV-2-infected lung | Non-infected Lung |

**Table S2. Details of the mouse models of COVID-19 datasets, Related to Figure 4.**

| | Present Data | GSE 154104 | GSE 166778 | GSE 174382 | GSE 180557 | GSE 189015 | GSE 205014 |
|---|---|---|---|---|---|---|---|
| **Up DEGs** | 3487 | 674 | 3145 | 4825 | 3219 | 1712 | 520 |
| **Dn DEGs** | 3174 | 305 | 2976 | 4204 | 2910 | 1318 | 174 |
| **Overlapped DEGs** | | | | | | | |
| **Present Data** | | 510 (14.0%) | 1203 (22.2%) | 1793 (27.5%) | 1690 (33.7%) | 837 (19.2%) | 423 (11.8%) |
| **GSE 154104** | 154 (4.6%) | | 388 (11.3%) | 475 (9.5%) | 445 (12.9%) | 347 (17.0%) | 259 (27.7%) |
| **GSE 166778** | 817 (15.3%) | 136 (4.3%) | | 1457 (22.4%) | 929 (17.1%) | 936 (23.9%) | 316 (9.4%) |
| **GSE 174382** | 1361 (22.6%) | 146 (3.3%) | 1374 (23.7%) | | 1433 (21.7%) | 1240 (23.4%) | 343 (6.9%) |
| **GSE 180557** | 1471 (31.9%) | 156 (5.1%) | 746 (14.5%) | 1370 (23.9%) | | 785 (18.9%) | 365 (10.8%) |
| **GSE 189015** | 372 (9.0%) | 46 (2.9%) | 635 (17.4%) | 782 (16.5%) | 458 (12.1%) | | 290 (14.9%) |
| **GSE 205014** | 96 (3.0%) | 13 (2.8%) | 40 (1.3%) | 55 (1.3%) | 55 (1.8%) | 31 (2.1%) | |

**Table S3. Overlap of differentially expressed genes (DEGs) among mouse COVID-19 models, Related to Figure 4.**
